# The contribution of silencer variants to human diseases

**DOI:** 10.1101/2024.05.17.24307558

**Authors:** Di Huang, Ivan Ovcharenko

## Abstract

Although disease-causal genetic variants have been found within silencer sequences, we still lack a comprehensive analysis of the association of silencers with diseases. Here, we profiled GWAS variants in 2.8 million candidate silencers across 97 human samples derived from a diverse panel of tissues and developmental time points, using deep learning models.

We show that candidate silencers exhibit strong enrichment in disease-associated variants, and several diseases display a much stronger association with silencer variants than enhancer variants. Close to 52% of candidate silencers cluster, forming silencer-rich loci, and, in the loci of Parkinson’s-disease-hallmark genes TRIM31 and MAL, the associated SNPs densely populate clustered candidate silencers rather than enhancers displaying an overall 2-fold enrichment in silencers versus enhancers. The disruption of apoptosis in neuronal cells is associated with both schizophrenia and bipolar disorder and can largely be attributed to variants within candidate silencers. Our model permits a mechanistic explanation of causative SNP effects by identifying altered binding of tissue-specific repressors and activators, validated with a 70% of directional concordance using SNP-SELEX. Narrowing the focus of the analysis to individual silencer variants, experimental data confirms the role of the rs62055708 SNP in Parkinson’s disease, rs2535629 in schizophrenia, and rs6207121 in Type 1 diabetes.

In summary, our results indicate that advances in deep learning models for discovery of disease-causal variants within candidate silencers effectively ‘double’ the number of functionally characterized GWAS variants. This provides a basis for explaining mechanisms of action and designing novel diagnostics and therapeutics.

## Introduction

A common but often elusive goal of biological investigations is to uncover the genetic basis of disease phenotypes (Zhang and Lupski 2015; Claussnitzer et al. 2020). This is challenging due to the inherent complexity of human genetics. Although genome-wide association studies (GWASs) offer valuable genetic insights into diseases and disorders, they struggle to pinpoint causative variants due to linkage disequilibrium among genetic variants. Notably, a significant majority of GWAS variants, exceeding 90%, occur within noncoding genomic regions (Watanabe et al. 2019). To accurately map disease-causing variants, it is vital to characterize the function of non-coding regions. Up to now, the investigations have primarily focused on well-characterized non-coding regulatory elements including enhancers, promoters, and insulators (Maurano et al. 2012; Farh et al. 2015; Finucane et al. 2015; Fulco et al. 2019; Konrad et al. 2019). These studies consistently underscore the impact of regulatory elements on disease susceptibility.

Evidence has also indicated pathological roles of silencers, however. For instance, a rare silencer variant disrupts binding of NR2F1 and affects the expression of *GATA2* in neurons leading to hereditary congenital facial paresis type 1 (Tenney et al. 2023). Another variant deactivates a silencer in breast cells, causing the overexpression of *ESR1* and *RMND1* in breast cancer (Dunning et al. 2016).

Despite these and a few similar discoveries, silencers have been underexplored in genetic and genomic research, in general, primarily due to the difficulties in systematically profiling these elements across the whole genome (Della Rosa and Spivakov 2020). Recent advancements in massively parallel reporter assays (MPRAs) and computational analysis tools have allowed genome-wide mapping of silencers (Doni Jayavelu et al. 2020; Pang and Snyder 2020; Huang and Ovcharenko 2022; Hussain et al. 2023; Zhu et al. 2023), opening doors to in-depth investigations into the association of silencers with diseases and phenotypic traits in humans.

Understanding the regulatory effects of non-coding variants is a key challenge in genetic research, essential to discovering molecular causes of diseases (Zhou et al. 2018; Li et al. 2023). Here, we apply a deep learning framework to a diverse collection of 97 biological samples (biosamples), building a deep learning model in each biosample to detect biosample-specific candidate silencers. Our results demonstrate that candidate silencers are enriched in disease-associated regulatory single-nucleotide polymorphisms (SNPs), but their disease-association profiles differ from those of enhancers. We demonstrate how silencer modeling can be used to predict the regulatory impact of variants within candidate silencers and to identify disease-causal variants.

## Results

### Genome-wide silencer landscape in 97 cell types

We trained two-phase deep learning TREDNet models (Hudaiberdiev et al. 2023) to predict enhancers and silencers, building a multi-class classifier for each of the biosamples collected by the ENCODE project (see Methods). Albeit lower than the 0.96 enhancer AUROC, the accuracy of silencer prediction was on par with our prior models (0.84 area under receiver operating characteristic curve, AUROC) (Huang and Ovcharenko 2022), and was significantly better than AUROC = 0.77 of our prior SVM model (Huang et al. 2019). While the SVM model employs DNA sequences and gene expression profiles for silencer prediction, the TREDnet model is DNA sequence-based, and thus can be readily extended to additional biosamples. These AUROC values exhibit a positive correlation with GC content levels and a negative correlation with repeat density (Figure 1A). This partially explains lower classification performance on silencers than on enhancers since enhancer sequences (defined as DNase-seq and H3K27ac ChIP-seq peaks) generally feature higher GC content and lower repeat density than silencers (defined as DNase-seq and H3K27me3 ChIP-seq peaks that lack overlap with H3K27ac peaks, see Methods). With the trained TREDNet models, we identified enhancers and silencers in each biosample, and conservatively selected 97 biosamples with over 5000 candidate enhancer and silencers in them for further investigation (Table S1). These biosamples encompass a diverse array of human cell types, including but not limited to 21 immune biosamples (19 blood cells, spleen and thymus), 17 digestive, metabolic and endocrine biosamples, and 6 biosamples from the central nervous system (Figure 1B and Table S1). Among them, 20 (20%) biosamples are from cancer cell lines.

**Figure 1.**
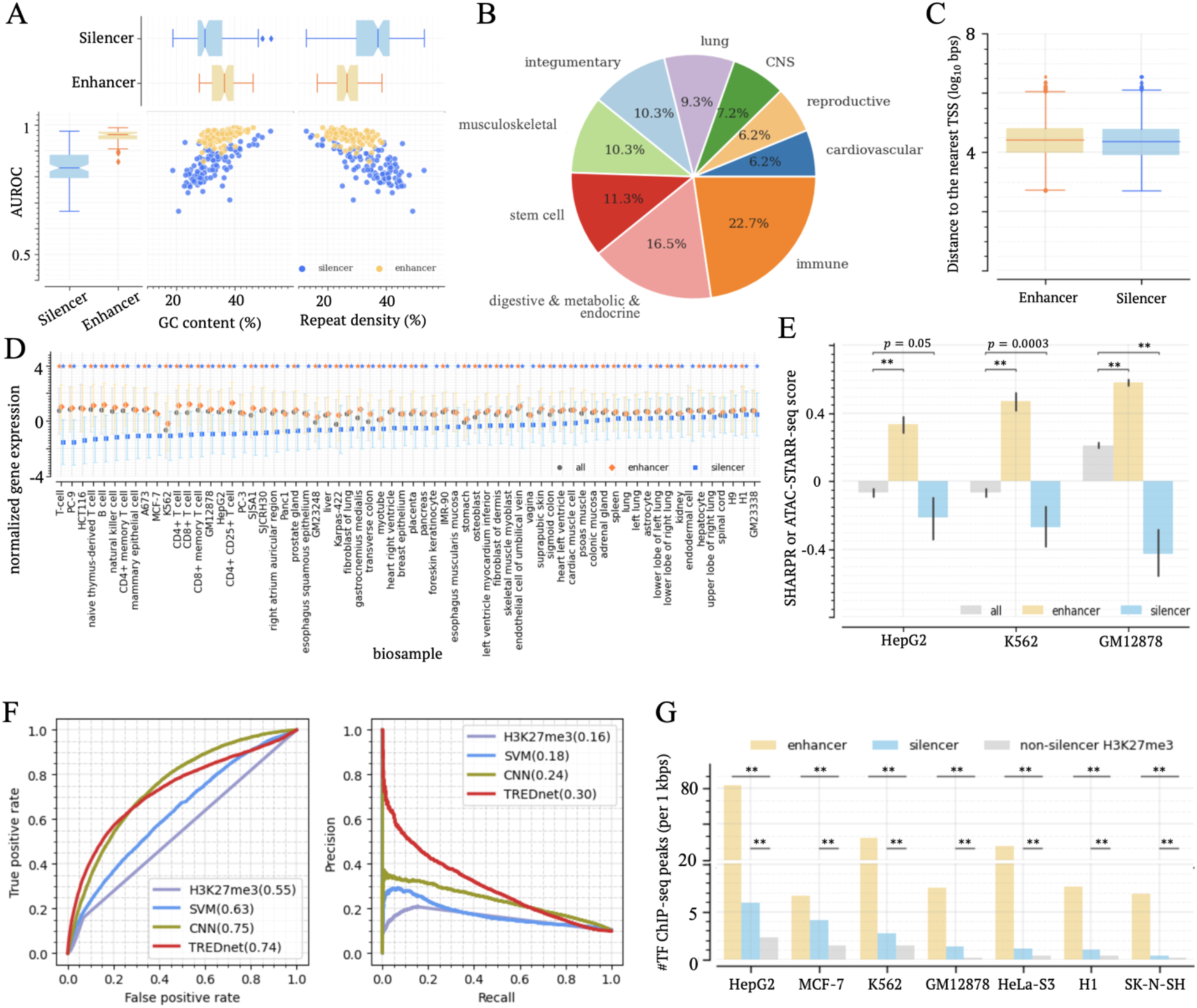
Profiling candidate silencers across 97 biosamples from diverse origins. (A) Classification performance (AUROCs) of TREDNet models for silencers and enhancers in analyzed biosamples. AUROCs exhibit correlation with GC contents and repeat densities of training sequence sets. Each dot represents a set of enhancers or silencers. (B) Distribution of 97 biosamples across groups. (C) Distance of candidate silencers and enhancers to their nearest TSSs. (D) Expression of genes proximal to candidate silencers and enhancers. Markers and their flanking lines represent the medians and standard deviations of gene expression levels. Blue and orange asterisks on the top represent the significantly low and high expression levels, respectively, compared to all genes (p < 0.05). (E) MPRA scores of candidate silencers and enhancers in three biosamples. (F) Performance of the TREDnet model on MPRA silencers. (G) Densities of TF ChIP-seq peaks within candidate silencers and enhancers across biosamples. ∗∗: p < 10^−10^

We identified a total of 2.8 million candidate silencers and 5.8 million enhancers (Supplementary Data 1), collectively spanning approximately 37.6% of the human genome. In cancer biosamples, 10% exhibit a higher count of silencers than enhancers, a proportion notably lower than the 17.9% of all examined biosamples (binomial test *p* = 0.007, Figure S1). This finding is consistent with gene overexpression in cancer cells (Santarius et al. 2010), which might be due to silencer loss or deactivation in cancer. On average, 57.7% of candidate silencers and 42.9% of candidate enhancers are located within intergenic regions (binomial test *p* < 10^−10^, Figure S2). Nonetheless, silencers and enhancers exhibit comparable distances to their nearest transcriptional start site (TSS), with approximately half of them residing within 26 kb of their nearest TSSs (Figure 1C).

Examining the evolutionary conservation, we noticed that an average of 8.7% of candidate silencer sequences and 10.6% of enhancer sequences overlap genomic regions conserved across 30 primate species (Siepel et al. 2005), significantly exceeding the 5.7% expectation stemming from the whole human genome (Student’s t-test *p* < 10^−10^, Figure S3). This underscores the negative selective pressure imposed on functional genomic regions to preserve their biological function (Siepel et al. 2005), but also reflects a rapid turnaround of regulatory elements in vertebrates (Villar et al. 2015). In 63.6% (15/22) of immune biosamples, candidate silencers are more conserved than enhancers, significantly higher than the 34% of all biosamples (binomial test *p* = 3 × 10^−9^). This finding highlights the significance of candidate silencers in immunological context. For example, the loci of *PCDHA/G* genes, which is highly conserved in vertebrates (Yu et al. 2007) and plays an important role in epithelial barrier formation and repair, displays the enrichment in candidate silencers, but not enhancers, in immune biosamples (Figure S4). The trend is also evident in the highly conserved loci of *HOXA* and *HOXD* clusters (Figure S4), developmentally essential genes associated with embryonic development (Quinonez and Innis 2014).

### Functional evaluation of silencer predictions

To assess the impact of candidate silencers, we initially analyzed the expression of genes located near these elements across 66 biosamples with available gene expression profiles from the ENCODE project (see Supplementary Notes) since genes associated with active silencers are likely to be lowly expression. Across all examined biosamples, genes neighboring candidate silencers exhibit significantly lower expression than all assayed genes (*p* < 0.05, Figure 1D). Similarly, genes targeted by candidate silencers, as determined by Hi-C chromatin loops (Salameh et al. 2020), consistently display notably low expression across all tested biosamples (*p* < 0.05, Figure S5).

Furthermore, we directly evaluated the activity of candidate silencers by utilizing the experimental results from MPRA platforms designed to measure silencing or activating impact of genomic regions. In K562 and HepG2 biosamples, candidate silencers frequently exhibit negative scores reported by the Sharpr-MPRAs (Ernst et al. 2016). These scores are significantly lower than those observed in enhancers and all tested regions (Wilcoxon rank-sum test *p* ≤ 0.05, Figure 1E), supporting the active silencing function of candidate silencers. Similarly, in GM12878, significant negative ATAC-STARR-seq scores, which represent “silent” genomic sequences (Hansen and Hodges 2022), are enriched among candidate silencers (*p* = 4 × 10^−16^ vs all tested sequences, Figure 1E).

Additionally, we compiled 7,701 K562 silencers from two independent MPRA studies based on ReSU (Pang and Snyder 2020) and STARR-seq (Doni Jayavelu et al. 2020). Of them, 541 overlap with K562 predicted silencers, which represents a significant enrichment compared to the DNase-seq peaks randomly selected from alternative biosamples and H3K27me3 ChIP-seq peaks not predicted as silencers in K562 (binomial test *p* < 10^−10^, Figure S6A). Similarly, in HepG2, predicted silencers are significantly enriched with silencers detected by the ReSU MPRA (Pang and Snyder 2020) (*p* < 10^−10^, Figure S6B).

Moreover, we validated the TREDnet silencer model on an independent experimental dataset of MPRA silencers. After excluding MPRA silencers overlapping sequences used for training the TREDnet model, we had 6,999 K562 MPRA silencers remaining for validation. On this subset of MPRA silencers, the TREDnet model demonstrates a classification performance of AUROC = 0.74 and AUPRC = 0.30 with the 1:9 ratio of positive to control samples. It shows a marginal improvement over our prior CNN classifier (Huang and Ovcharenko 2022) and significantly outperforms our prior SVM model (Huang et al. 2019), and general H3K27me3 signal profiles (Figure 1G). Furthermore, the TREDnet silencer model can effectively distinguish both H3K27me3 and non-H3K27me3 MPRA silencers from control sequences (Figure S7). These results reaffirm that the TREDnet silencer model can identify active silencers with respectable accuracy.

To further investigate whether candidate silencers actively suppress gene expression as opposed to being genomic regions of repressed chromatin, we analyzed the abundance of transcription factor binding sites (TFBSs) and chromatin contacts, under the assumption that repressed chromatin regions host significantly fewer TFBSs and chromatin contacts than active enhancer and silencer regions. In each of tested biosample with ChIP-seq data for more than 50 TFs available from the ENCODE project, candidate silencers contain, on average, 3.5 times as many TF ChIP-seq peaks as H3K27me3 ChIP-seq peaks lacking candidate silencers (Wilcoxon rank-sum test *p* < 10^−10^, Figure 1E). Additionally, the density of Hi-C chromatin contacts within predicted silences is 1.5 times greater than the corresponding density within H3K27me3 ChIP-seq peaks lacking candidate silencers (binomial test *p* < 0.05, Figure S8).

Overall, these results support that the TREDnet predicted silencers predominantly act as active silencers and not simply heterochromatic regions of the genome. Therefore, we refer to them as candidate silencers.

### Candidate silencers are associated with development

To evaluate biological functions associated with candidate silencers, we turned to their nearby genes. Genomic proximity to a specific class of genes, although not comprehensive enough to capture long-range chromatin interactions, are commonly used to examine biological functions of regulatory elements (McLean et al. 2010). We defined the locus of a gene as its gene body along with the entire intergenic areas between this gene and its nearest neighbors. On average, 6.3% of gene loci are enriched in candidate silencers with a significancy of *p* < 10^−5^ compared to the whole genome (referred to as “silencer-rich gene loci”). This percentage is substantially higher than the 4.7% of gene loci showing enhancer enrichment (Student’s *t*-test *p* = 0.0007, Figure 2A). Across biosamples, silencer-rich loci harbor 51.7% of all silencers, notably higher than the 25.8% of enhancers found in enhancer-rich loci (Student’s *t*-test *p* = 2 × 10^−22^, Figure 2A), suggesting a pronounced trend of candidate silencer accumulation in specific gene loci.

**Figure 2.**
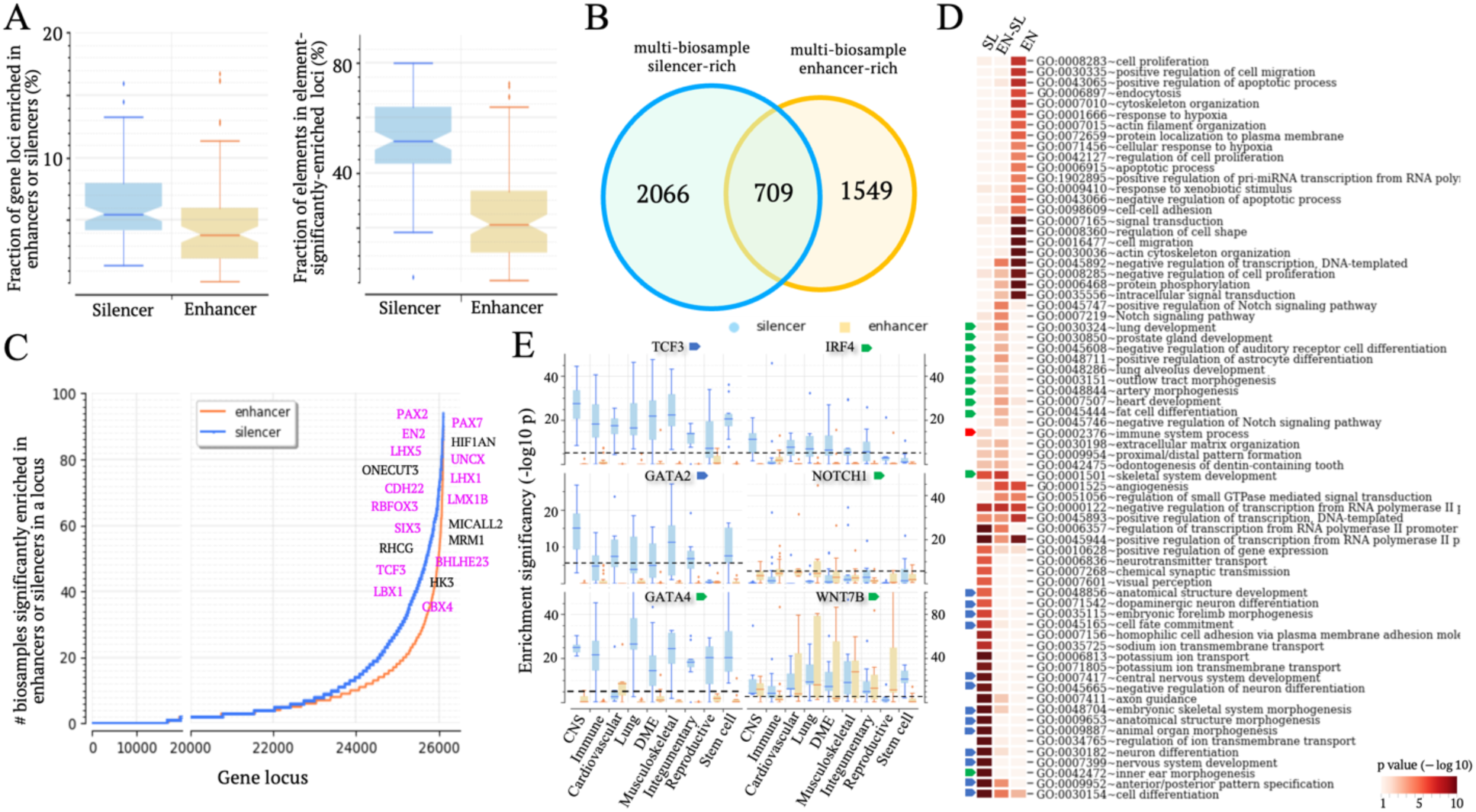
Candidate silencers are significantly associated with development and immunity. (A) Fractions of silencer-rich or enhancer-rich gene loci (the left panel). Proportions of candidate silencers located within silencer-rich loci and enhancers located within enhancer-rich loci are shown in the right panel. (B) Numbers of multi-biosample silencer-rich and enhancer-rich gene loci. Notably, 709 gene loci are both multi-biosample silencer-rich and enhancer-rich. (C) Frequency of gene loci exhibiting silencer-rich (blue line) or enhancer-rich (the orange line) across biosamples. Top-frequency silencer-rich gene loci are listed, among which developmental loci are highlighted in pink. (D) Heatmap illustrating biological processes significantly associated with different gene sets. SL and EN represents multi-biosample silencer-rich and enhancer-rich gene loci, respectively. ENSL represents the intersection of SL and EN sets. Biological processes in embryonic and central nervous system (CNS) development are indicated by blue arrows, while immunity regulation and tissue-specific development are by red and green arrows, respectively. (E) Enrichment of candidate silencers and enhancers in six gene loci. *The dash lines represent the threshold* (*p* = 1.9 × 10^−6^*) for significant enrichment (see Supplementary Notes). The upper and lower whisker edges in these boxplots represent approximately 25% and 75% quartiles of the presented data*.

Among the gene loci displaying the highest frequency of candidate silencer enrichment across biosamples are *PAX2*, *PAX7*, *EN2*, *HIF1AN*, *LHX5* (Figure 2B). All of them are known as essential for development. Gene loci significantly enriched in candidate silencers in over-9 biosamples from different groups are denoted as multi-biosample silencer-rich gene loci. In total, there are 2,775 such gene loci (Figure 2C). These genes are associated with fundamental developmental processes and neurological system development (DAVID hypergeometric test *p* < 10^−6^, indicated by blue arrows, Figure 2D) (Sherman et al. 2022). Additionally, these gene loci are notably associated with immune system regulation (*p* = 0.002). For example, the loci of cell-differentiation regulators *TCF3* and *GATA2* show elevated densities of candidate silencers in 90% and 54.6% of examined biosamples, respectively. The *IRF4* locus, crucial for the immune system, displays a significant enrichment in candidate silencers in 71.4% of CNS cells.

On the other hand, multi-biosample enhancer-rich gene loci are involved in housekeeping biological processes such as signal transduction, cell-cell adhesion, and protein phosphorylation (*p* < 10^−3^, Figure 2D). Furthermore, there are a total of 709 gene loci that are both multi-biosample enhancer-rich and silencer-rich, thus termed as multi-biosample enhancer-silencer-rich (Figure 2C). These genes often take part in tissue-specific developmental processes (indicated by green arrows in Figure 2D). For example, the locus of *GATA4*, a key factor in heart, pancreatic and hepatic development, is enhancer-rich in cardiovascular biosamples but silencer-rich in 50% of other biosamples (Figure 2E). The locus of *WNT7B*, encoding a signal protein crucial for tissue development, is silencer-rich in 79.4% of biosamples and enhancer-rich in 53.6% of them. In summary, candidate silencers are preferentially distributed in the proximity of the genes controlling fundamental and tissue-specific developmental processes, significantly associated with the regulation of these genes. These results suggest that the regulation of developmental genes is often tightly orchestrated with an array of enhancer and silencer elements establishing a complex multi-cellular regulatory profile.

### Silencer-to-enhancer transitions are a hallmark of cellular differentiations

Functional transitions between enhancers and silencers across biological contexts are pivotal in the precise and expeditious regulation of developmental processes (Erceg et al. 2017; Huang and Ovcharenko 2022). A substantial portion of candidate silencers and enhancers reported here have dual functions. Specifically, 55% of candidate silencers and 42% of enhancers are dual functional regulatory elements (DFREs), acting as enhancers in certain biosamples but as silencers in others (Figure S9).

Moreover, 68% of candidate silencers of H1 human embryonic stem cells (H1-hESCs) are converted to enhancers in partially or fully differentiated biosamples examined in this study. These enhancers contain significantly more TFBSs than other enhancers in five out of six tested biosamples (*p* < 10^−10^). This significance remains evident even when compared to the enhancers that are converted from H1-hESC poised enhancers (PEs, defined as H3K4me1 ChIP-seq peaks carrying no H3K27ac modification signals in H1-hESCs). For example, in K562 cells, each hESC-silencer-converted enhancer harbors an average of 58 TF ChIP-seq peaks, significantly more than the 35 found in all K562 enhancers and the 42 in K562 hESC-PE-converted enhancers (*p* < 10^−10^, Figure 3A). Moreover, compared to other enhancers (including PE-converted enhancers), hESC-silencer-converted enhancers are enriched in TF ChIP-seq peaks of dual functional TFs like YY1 and chromatin organizers such as CTCF, RAD21, and ZNF143. On the other hand, these enhancers lack TF ChIP-seq peaks of cell-specific transcriptional activators like CEBPB in HepG2 cells, ESR1 and NEUROD1 in MCF-7 cells, BACH1 and EBF1 in K562 cells, IRF4 and BCL11A in GM12878 cells (Figure 3B). Furthermore, in 94% (65/69) of the biosamples for which CTCF ChIP-seq data are available in the ENCODE project (Table S2), hESC-silencer-converted enhancers show significantly higher densities of CTCF ChIP-seq peaks compared to all enhancers, including hESC-PE-converted ones (Figure 3C), with an average enrichment fold of 1.8. The pronounced enrichment in TF ChIP-seq peaks, particularly for CTCF, hints that hESC-silencer-converted enhancers frequently serve as anchors for chromatin loops, a crucial aspect in chromatin organization (Clyde 2023).

**Figure 3.**
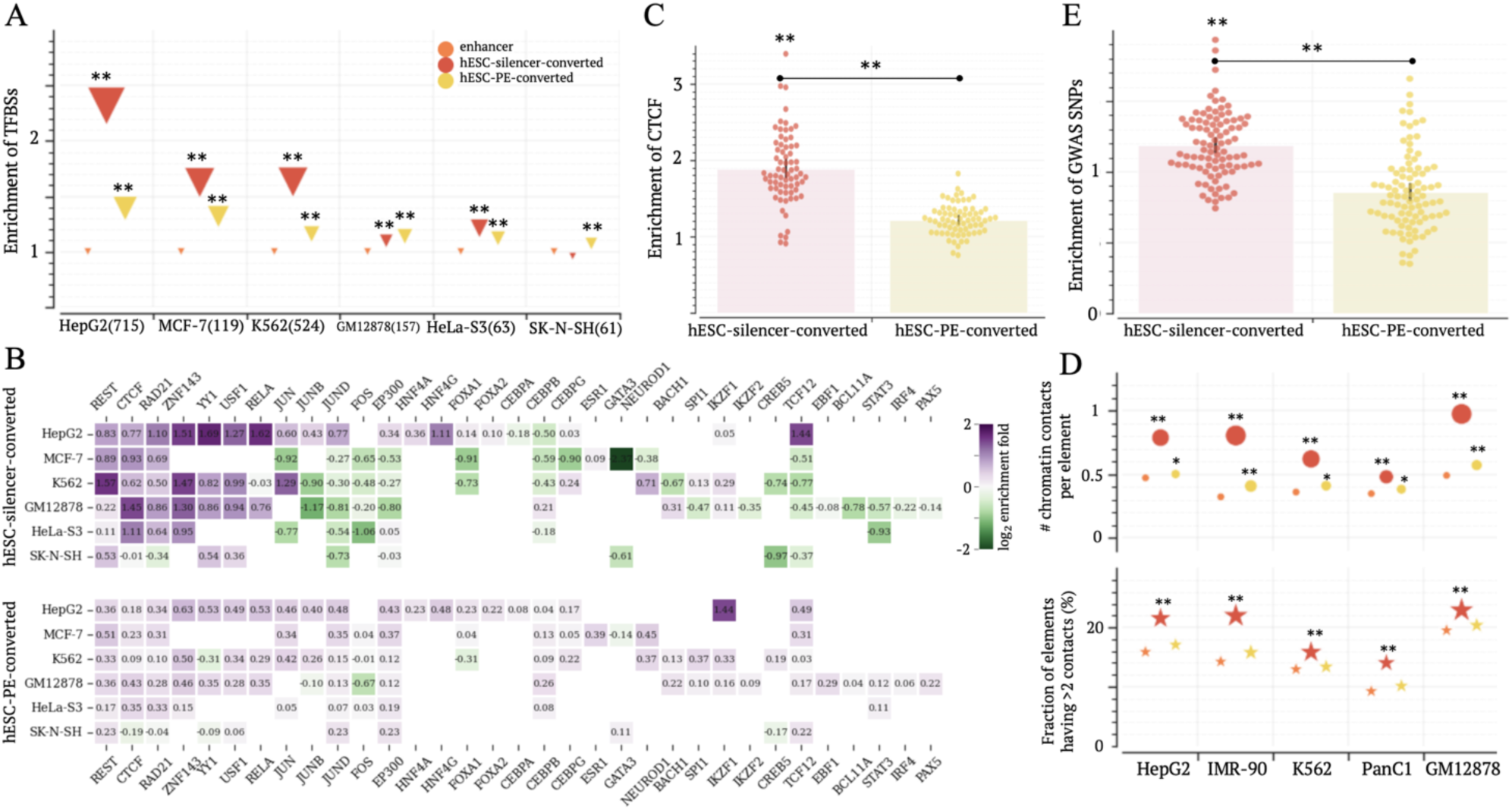
hESC-silencer-converted enhancers anchor chromatin loops. (A) Enrichment of ChIP-seq TFBSs in hESC-silencer-converted and hESC-PE-converted enhancers in comparison to all enhancers. The numbers in parentheses are the number of TFs examined in this study. (B) Enrichment of TFBSs for individual TFs. The blank cells indicate absence of TF ChIP-seq data. (C) Enrichment of CTCF ChIP-seq TFBSs across 69 biosamples. (D) Numbers of chromatin contacts per elements (the top panel) and the fractions of elements having >2 contacts (the bottom panel) in hESC-silencer-converted enhancers. Additional results are presented in Figure S2. (E) Enrichment of GWAS SNPs within hESC-silencer-converted and hESC-PE-converted enhancers in comparison to all enhancers across biosamples. ∗: *p* < 0.01 and ∗∗: *p* < 10^−10^.

To further verify this interpretation, we analyzed chromatin contacts of enhancers (as defined by Hi-C data, see Methods). In the biosamples where over 20% of enhancers have reported Hi-C contacts, hESC-silencer-converted enhancers display the highest density of Hi-C contacts (*p* < 10^−10^, Figure 3D). Importantly, they hold at-least-3 chromatin contacts more frequently than other enhancers (*p* < 10^−10^). In K562 cells, 14% of hESC-silencer-converted enhancers have at-least-3 chromatin contacts, significantly higher than the 9.2% of all enhancers and the 10.1% of hESC-PE-converted enhancers (*p* < 10^−10^, Figure 3D). These trends persist in biosamples where fewer than 20% of enhancers have Hi-C contacts, although statistical significance diminishes possibly due to limited detection of chromatin contacts (Figure S10). These results reaffirm that hESC-silencer-converted enhancers often serve as anchors for chromatin loops.

To further assess the functional significance of hESC-silencer-converted enhancers, we utilized the single nucleotide polymorphisms (SNPs) annotated in GWASs. We downloaded GWAS SNPs documented in the National Human Genome Research Institute (NHGRI) catalog (McMahon et al. 2018) and in UK Biobank release 2 cohort (Bycroft et al. 2018). After the inclusion of the SNPs in tight linkage disequilibrium (LD r^2^> 0.8) with GWAS SNPs, a total of 2.2 million GWAS SNPs were compiled, which are associated with 1,116 distinct traits (Figure S11, see Methods). HESC-silencer-converted enhancers exhibit a significant increase (*p* < 0.01) in the density of GWAS SNPs compared to all enhancers in 75% (69/92) of differentiated biosamples (Figure 3E). This increase remains significant even when compared to H1-hESC-PE converted enhancers (*p* < 10^−10^). In 73% (67/92) of differentiated biosamples, GWAS SNP densities in hESC-silencer-converted enhancers are significantly higher than those in hESC-PE-converted enhancers. These findings support the functional importance of these enhancers, partially due to their role as anchors for chromatin loops.

### GWAS studies suggest a critical role of candidate silencers in neurological and autoimmune disorders

We further utilized GWAS SNPs to assess the phenotypic impact of all candidate silencers. On average, candidate enhancers and silencers in examined biosamples harbor 3.4 and 3.0 NHGRI GWAS SNPs per 1kb, respectively. Both values are significantly higher than the 2.4 whole genome GWAS SNPs density (Student’s *t*-test *p* < 10^−20^, Figure 4A).

**Figure 4.**
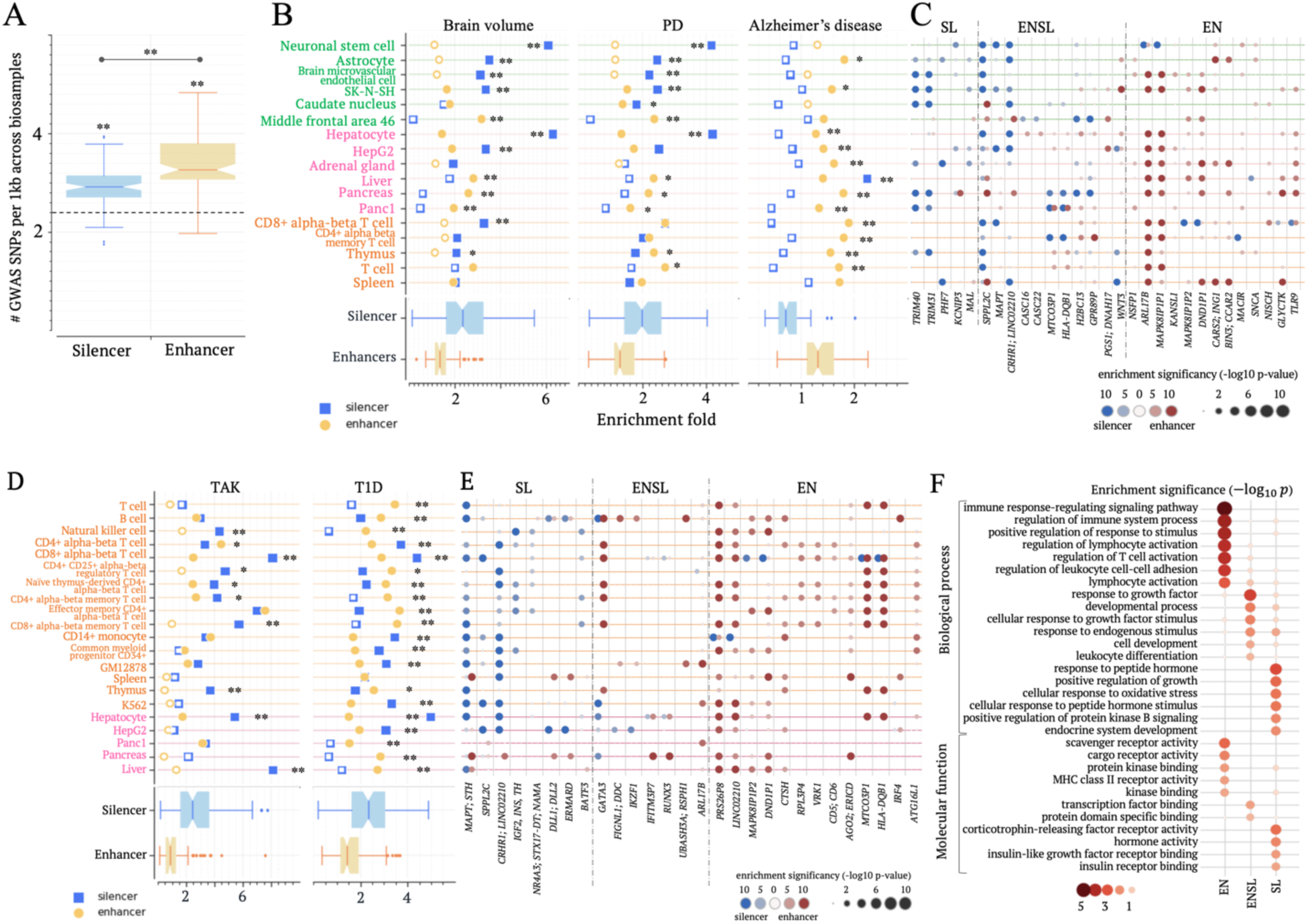
Candidate silencers exhibit the enrichment for GWAS SNPs. (A) Numbers of GWAS SNPs per 1kb in candidate silencers and enhancers across biosamples. The dash line represents the number of GWAS SNPs per 1kb in the whole genome. (B) Enrichments of SNPs associated with brain volume, PD and Alzheimer’s disease within candidate silencers and enhancers across biosamples. Asterisks indicate the significant difference between candidate silencers and enhancers. (C) Enrichments of PD-associated SNPs within candidate silencers and enhancers in individual gene loci. Only gene loci having significant enrichments are included here. (D) Enrichments of SNPs associated with TAK and T1D within candidate silencers and enhancers. In (B) and (D), enrichment folds are estimated in comparison to the whole genome. Significant enrichments are denoted by solid markers (*p* < 10^−5^). The results on other autoimmune diseases are presented in Figure S7. (E) Enrichment of T1D-associated SNPs within candidate silencers and enhancers in individual gene loci. In (C) and (E), gene loci are clustered based on the enrichment profiles of associated SNPs. SL/EN represent the gene loci where the associated SNPs are enriched exclusively in candidate silencers/enhancers, while ENSL denotes the gene loci where the associated SNPs are enriched in both candidate silencers and enhancers. (F) Functional analysis results for T1D-associated gene clusters defined in (E). ∗∗: *p* < 10^−5^ and ∗: *p* < 0.01.

Similarly, candidate silencers exhibit significant enrichment in expression quantitative trait loci (eQTLs) obtained from the GTEx project (The GTEx Consortium 2015) compared to the whole genome across 28 out of 40 examined biosamples (Figure S12A, see Supplementary Notes). Additionally, candidate silencer eQTLs achieve significance levels akin to enhancer eQTLs across these biosamples (Figure S12B). Silencer eQTLs are, however, more tissue-specific than enhancer eQTLs in 90% of examined biosamples (36/40; *p* < 0.05, Figure S12C). Furthermore, we explored the distribution of GWAS SNPs deposited to the ClinVar archive (Landrum et al. 2020). Candidate silencers host 1.47 ClinVar SNPs per 1 kb. This density exceeds 1.29 ClinVar SNP per 1 kb within enhancers, with both densities significantly surpassing the expected 0.76 ClinVar SNP per 1 kb baseline from the whole genome (*p* < 10^−5^, Figure S13A). We also examined the distribution of cancer somatic variants compiled in the ICGC database (Zhang et al. 2011). These cancer variants show significant enrichment within candidate silencers in the matched biosamples for seven out of eight examined cancers (Figure S13B). For example, the density of myeloid variants in K562 candidate silencers is 1.3 times that expected from the whole genome baseline. Taken together, these findings suggest an observable phenotypic impact of candidate silencers.

Notably, GWAS SNPs associated with different traits have varying enrichment levels in candidate silencers and enhancers across biosamples (Table S3). For example, SNPs associated with Alzheimer’s disease are predominantly located in CNS and immune system enhancers (*p* < 10^−10^ versus the whole genome as marked by a solid symbol, Figure 4B). In contrast, SNPs associated with Parkinson’s disease (PD) are preferentially located in candidate silencers in five out of six CNS biosamples (*p* < 10^−5^ versus the whole genome and enhancer counterparts) and within enhancers in immune biosamples (Figure 4B). SNPs associated with brain volume traits, such as intracranial, hippocampal, thalamus, and subiculum volume, are notably biased towards candidate silencers in five out of six CNS biosamples (*p* < 10^−5^ versus the whole genome and enhancer counterparts, Figure 4B).

To further dissect the genetic basis of PD, we evaluated the enrichment levels of associated SNPs within candidate silencers and enhancers in each gene locus (see Methods). In the locus of *TLR9,* a gene known for its involvement in the degeneration of dopamine neurons in PD (Maatouk et al. 2018), PD-associated SNPs mainly cluster in CNS enhancers (Figure 4C). In contrast, the *TRIM31* locus, responsible for metal ion binding, harbors a total of 104 PD-associated SNPs, a number significantly higher than the genome-wide average (*p* < 10^−30^). Of these SNPs, 18 are located within SK-N-SH candidate silencers, which is notably higher than 8 SNPs as expected in the TRIM31 locus. Interestingly, no PD-associated SNPs are found within the *TRIM31* SK-N-SH candidate enhancer. This pronounced bias to CNS candidate silencers is also observed in the loci of *MAL* and *MAPT*, both associated with neurogenesis (Figure 4C). These findings consistently underscore the significant role of CNS candidate silencers in PD, particularly in relation to metal ion binding and neurogenesis, two factors closely linked to PD (Figure S14) (Marxreiter et al. 2013; Moons et al. 2020).

We also analyzed the genetic mechanisms underlying differences in brain volume. The SNPs associated with brain volume are enriched within candidate enhancers in the loci of *CTBP2* and *ZRANB1* in CNS biosamples and *KANSL1* in immune biosamples. These SNPs are enriched in candidate silencers in the locus of *DMRAT2* in CNS biosamples (Figure S15). *DMRTA2* is key in controlling the cell cycle during neuronal differentiation. Its dysregulation may lead to severe microcephaly (Young et al. 2017), suggesting the crucial contribution of CNS candidate silencers to brain volume measurement and, more broadly, the development of CNS.

Similarly, across autoimmune disorders, candidate enhancers and silencers in immune and endocrine biosamples show varying enrichments for GWAS SNPs. For example, while enriched within both candidate enhancers and silencers (*p* < 10^−5^ vs the whole genome), SNPs associated with rheumatoid and system lupus erythematosus (SLE) exhibit a distinct predilection for immune enhancers but for endocrine candidate silencers (silencers vs enhancers: *p* < 10^−5^, Figure S16). On the other hand, osteoarthritis-associated SNPs prefer candidate silencers over enhancers in immune system biosamples (silencers vs enhancers: 2.3 vs 2.0 of the average enrichment, binomial test *p* = 10^−20^). Takayasu’s arteritis (TAK) associated SNPs are preferentially situated within candidate silencers in immune system biosamples (silencers vs enhancers: 3.9 vs 2.4 of the average enrichment, *p* = 10^−11^, Figure 4D). Especially, in the MICA locus, TAK-associated SNPs are clustered within candidate silencers, rather than enhancers, in immune system biosamples (Figure S17). Given that the upregulation of MIC family in blood vessels contributes to the stimulation of natural killer cells in TAK (Yoshifuji and Terao 2020), it is plausible that the deactivation of candidate silencers in immune system biosamples could underlie the etiology of TAK.

Interestingly, SNPs associated with type 1 diabetes (T1D), a T-cell-mediated autoimmune disease that attacks pancreatic β cells (Steck and Rewers 2011), are notably prevalent within both candidate silencers and enhancers across immune system and endocrine biosamples (Figure 4D). However, these SNPs display varying preferences for candidate silencers and enhancers within individual gene loci (Figure 4E). Gene loci enriched with T1D-associated enhancer SNPs govern immune processes and/or the activity of receptors (Figure 4F). Instances include *IRF4*, *CD5*, *CD6* and *CTSH*. In contrast, T1D-associated silencer SNPs congregate conspicuously within the loci of *INS*, *IGF2*, and several other genes responsive to or producing hormones, notably insulin. Overexpression of *IGF2* renders pancreas islets susceptible to immune onslaught, thereby potentially serving as a key biomarker of T1D pathogenesis (Casellas et al. 2015). Our finding proposes that silencer variants in *IGF2* locus may contribute to T1D risk and identify a handful of specific silencer SNPs, which could be targeted in follow-up clinical and biochemical studies.

In short, candidate silencers and enhances, thought governing distinct functions, jointly drive crucial biological progress in complex diseases, as exemplified here by PD, T1D and TAK. However, silencers’ contributions to these diseases are not identical to those of enhancers.

### Candidate silencers underly the genetic difference between bipolar disorder and schizophrenia

To demonstrate the application of candidate silencer (and enhancer) profiles in a disease genetic study, we investigated regulatory mechanisms of bipolar disorder (BPD) and schizophrenia (SCZ). These two neurodevelopmental disorders, with a genetic correlation of over 0.6 based on common SNPs (Lee et al. 2013), share substantial overlap in both genetics and symptomology. The identification of shared and distinct genetic components between SCZ and BPD constitutes a fundamental stride toward deciphering the mechanisms of these diseases and formulating targeted therapeutic interventions (Ruderfer et al. 2018). To address this objective, we utilized candidate silencer and enhancer profiles in CNS, immune system and endocrine biosamples, given the notable involvement of endocrine and immune systems in these disorders (Kemp et al. 2010; Severance et al. 2020). SNPs associated with SCZ and/or BPD are enriched in candidate silencers and enhancers across endocrine and immune biosamples (Figure 5A). SCZ-associated SNPs are enriched in CNS candidate enhancers, while BPD-associated SNPs are preferentially distributed within CNS candidate silencers (*p* < 0.001 vs the whole genome, Figure. 5A).

**Figure 5.**
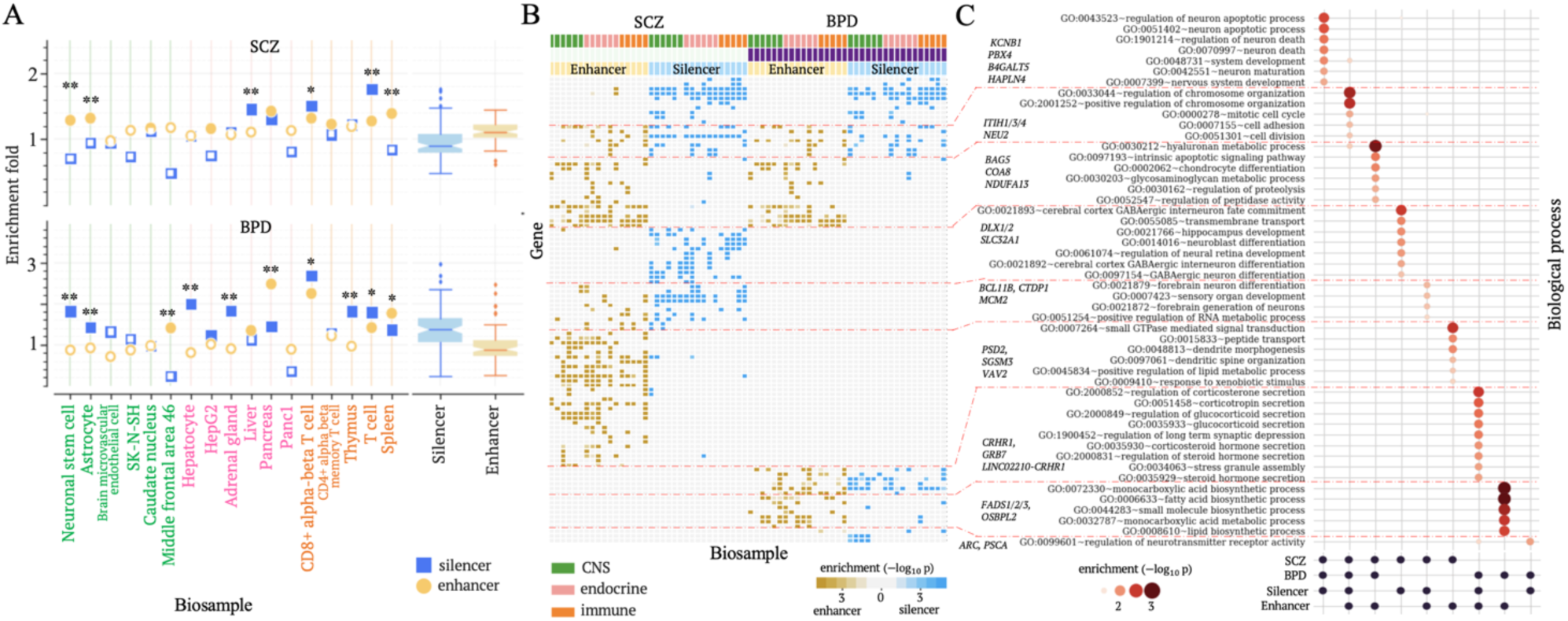
Candidate silencers distinguish SCZ from BPD. (A) Enrichments of SNPs associated with BPD and SCZ within candidate silencers and enhancers across biosamples. Asterisks by the markers indicate the significant difference between candidate silencers and enhancers. (B) Heatmap depicting the clusters of gene loci associated with SCZ and/or BPD, based on the enrichment profiles of associated SNPs within candidate silencers and enhancers. Each column represents the enrichment of SCZ or BPD associated SNPs within candidate silencers or enhancers in a biosample. The biosamples presented here are the same as those in (A). (C) Functional analysis of gene clusters defined in (B). ∗: *p* < 0.001 and ∗∗: *p* < 10^−5^.

To further elucidate genetic factors contributing to SCZ and BPD, we analyzed the distribution of their associated SNPs in each gene locus. Both SCZ– and BPD-associated SNPs display enrichment within enhancers in the loci of genes responsible for housekeeping biological activities like intrinsic apoptosis and hyaluronan metabolic process (*p* < 0.01, Figure 5B). In contrast, these SNPs are commonly found within candidate silencers in the loci of CNS-specific genes, particularly those controlling the apoptosis of neuronal cells and CNS development. For example, the locus of *KCNB1*, a key gene in the voltage-gated potassium channel crucial for neuron development and apoptosis (Bortolami et al. 2023), harbors 38 SCZ-associated SNPs and 21 BDP-associated SNPs. These numbers significantly exceed the expected by chance from the whole genome (*p* < 10^−22^). Among 38 SCZ-associated SNPs in the *KCNB1* locus, 10 (21.1%) are located within astrocyte candidate silencers, a notable preference as compared to the mere 1.2% of all SCZ-associated SNPs found in astrocyte candidate enhancers (binomial test *p* = 10^−11^). Similarly, in the *KCNB1* locus, 8 (38.1%) of the BDP-associated SNPs are located within astrocyte candidate silencers, significantly higher than the 2.9% observed for all BPD-associated SNPs across the whole genome (binomial test *p* = 10^−7^). The significant association of SCZ and BDP with neuron development and apoptosis, consistent with the previous findings (Benes 2004; Clifton et al. 2019), emphasizes the crucial role of silencer variants in the susceptibility to BPD and SCZ (Figure 5B).

Interestingly, despite an insignificant enrichment in CNS candidate silencers on a genome-wide level, SCZ-associated SNPs exhibit a distinct enrichment within candidate silencers in the loci of genes controlling the differentiation of GABAergic interneuron cells and hippocampus development (Figure 5C). Aberrant activity of GABAergic neurons has been reported as a key site of SCZ pathology (Jahangir et al. 2021). Our finding proposes that this anomaly is greatly attributable to the variants in CNS candidate silencers, thereby offering a lead for further biological examinations.

On the other hand, BPD-associated SNPs are enriched within both candidate silencers and enhancers in the loci of genes regulating corticosterone secretion and long-term synaptic depression. These two biological processes have been observed to be dysregulated in BPD patients (Du et al. 2011; Faurholt-Jepsen et al. 2021). In summary, analyzing candidate silencer and enhancer profiles alongside GWAS results can unveil the biological mechanisms that differentiate diseases with similar origins, as demonstrated by the analysis of BPD and SCZ here.

### Disease-associated silencer variants alter binding affinities of TFs

Our investigation next proceeded to the analysis of individual SNPs, aiming to identify disease-causal or trait-determining non-coding variants among GWAS SNPs (Zhou et al. 2018; Huang and Ovcharenko 2022). We quantified the impact of SNPs on gene regulation by comparing prediction scores from a trained TREDNet model between SNP alleles, denoted as Δ*repression* (see Methods). A positive Δ*repression* suggests a decrease in repressive activity due to a given SNP. SNPs with a significant Δ*repression* are marked as regulatory-activity-alternating SNPs (raSNPs, see Methods). RaSNPs are more frequently found in TF ChIP-seq peaks than common SNPs across seven biosamples (binomial test *p* < 10^−10^, Figure 6A). To prevent possible bias of raSNPs towards specific TFs, all seven biosamples examined in this study include ChIP-seq peaks for more than 50 TFs (see Method). In HepG2, a candidate-silencer raSNP coincides with an average of 2.1 TF ChIP-seq peaks, which is 1.22 times the average for all common SNPs within candidate silencers (*p* < 10^−10^, Figure 6A). Similarly, in enhancers, TF ChIP-seq peak densities at raSNPs are 1.33 times those at all common SNPs (*p* < 10^−10^).

**Figure 6.**
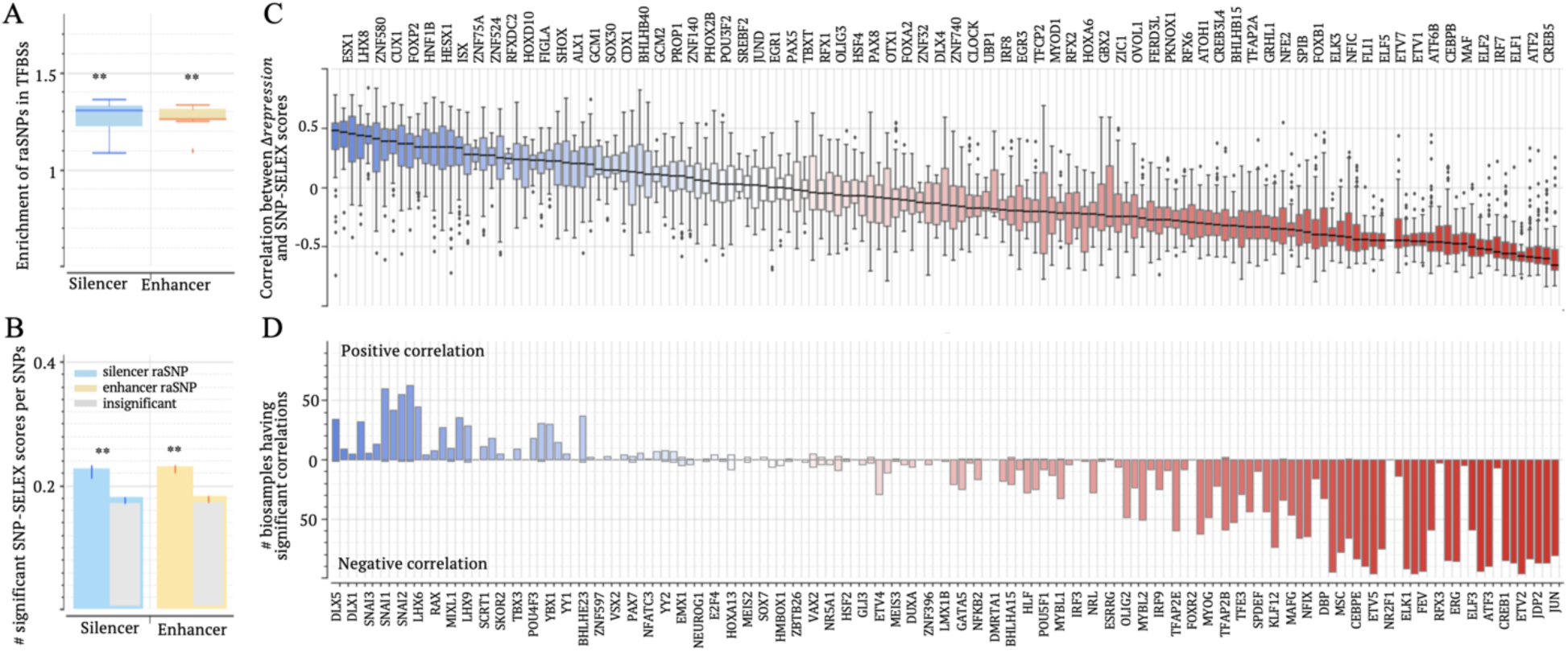
Δ*repression* significantly correlates with SNP-SELEX scores. (A) Enrichments of raSNPs in TFBSs (as defined in TF ChIP-seq peaks, Table S2) in seven biosamples. (B) Enrichments of significant SNP-SELEX scores among raSNPs. (C) Correlations between Δ*repression* and SNP-SELEX scores for each TF across biosamples. (D) Frequency distribution of biosamples exhibiting significantly positive (above zero line) and negative (below zeros line) correlations between Δ*repression* and SNP-SELEX scores for each TF. TF names are displayed along the bottom x-axis in (C) and the bottom x-axis in (D) combined. ∗∗: *p* < 10^−10^.

We then evaluated allele-specific TF-binding affinities of raSNPs. Allele-specific TF-binding affinities of SNPs were measured in a multiplex protein-DNA binding assay, known as systematic evolution of ligands by exponential enrichment (SNP-SELEX), for 270 TFs in the HepG2 cell line (Yan et al. 2021). Significant SNP-SELEX scores, which indicate substantial difference in binding affinities between SNP alleles, frequently occur among raSNPs across all examined biosamples. The occurrence rates of significant SNP-SELEX scores at raSNPs are over 1.26 times those at SNPs with insignificant-Δ*repression* scores, within either candidate silencers or enhancers (binomial test *p* < 10^−10^, Figure 6B, see Methods). These high occurrence frequencies, together with the enrichment of raSNPs in TF ChIP-seq peaks, highlight the significant possibility of raSNPs altering TF binding affinities.

Importantly, Δ*repression* scores positively correlate with SNP-SELEX scores of transcription repressors. For the repressors FOXP1 and SNAI1/2 (The Alliance of Genome Resources 2020), these positive correlations are significant (linear regression *p* < 0.05) in over-50 biosamples (Figures 6C and 6D). Of the raSNPs having significant SNP-SELEX scores for FOXP1, 69% show the directional concordance between Δ*repression* and SNP-SELEX scores (Figure S18). This concordance rate is over 65% for SNAI1/2. In contrast, Δ*repression* scores negatively correlate with SNP-SELEX scores of transcription activators. For prominent activators like JUN, CREB5, ELF1/2, CEBPE, NFE2 and SPIB, these negative correlations remain significant in over-50 biosamples. On average, the directional discordance rates between Δ*repression* and SNP-SELEX scores for these TFs is 67%. As positive SNP-SELEX scores indicate a reduction in binding affinity from wild-type to mutant alleles, these substantial positive or negative correlations (and directional concordance or discordance rates) underscore the effectiveness of Δ*repression* scores in capturing the impact of SNPs on binding affinity for both transcriptional repressors and activators. Additionally, bifunctional TFs like YY2 and PAX5, which act as both activators and repressors, rarely present a significant Δ*repression*-SNP-SELEX correlation in examined biosamples (Figures 6C, 6D and S18).

For example, the SNP rs11065189, associated with SCZ but not BPD, is situated within a candidate silencer in brain microvascular endothelial cells. The substitution from G to A results in a significant decrease in binding affinity of the transcriptional activators MAF, MAFG and NRL. These measurements align with Δ*repression* = −0.49, the highest magnitude within its 5kb vicinity (Figure S19).

In summary, these three lines of TF-binding-based evidence consistently substantiate the functional potency of raSNPs and the accuracy of Δ*repression* scores in evaluating the influence of SNPs on TF-binding affinity.

### The role of silencer SNPs in PD, SCZ and other neurological diseases

To directly evaluate the relationship between Δ*repression* scores and raSNPs, we resorted to the outcomes of MPRA experiments that assess allele-specific impacts of SNPs on gene regulation. Although these MPRA platforms were not specifically tailored for silencer SNPs, they provide valuable insights. For example, in SuRE MPRA experiments conducted in K562 cells (van Arensbergen et al. 2019), 19,237 SNPs were reported to significantly alter regulatory activity, known as reporter assay QLTs (raQTLs). These raQLTs are extremely enriched in K562 enhancers, consistent with previous findings (van Arensbergen et al. 2019). Nevertheless, we also observed a significant enrichment of raQTLs in candidate silencers and K562 MPRA silencers compared to the whole genome and H3K27me3 ChIP-seq peaks not classified as silencers (binomial test *p* < 10^−10^), although these silencer enrichment levels are notably lower than that in enhancers (*p* < 10^−10^, Figure S20A), as expected from the nature of the experimental data. This enrichment further supports the active state of K562 candidate silencers. In addition, Δ*repression*s are positively correlated with raQLT scores, irrespective of whether these raQTLs are in silencers or enhancers (Figure S20B). Taken together, MPRA scores by which the difference in regulatory influence between SNP alleles are quantified, though not specifically designed for silencer SNPs, can be used to examine the performance of Δ*repression*s in prioritizing disease-risk SNPs within candidate silencers.

To directly evaluate the regulatory impacts of raSNPs in candidate silencers in CNS biosamples, we utilized their MPRA scores on dementia GWAS SNPs (Cooper et al. 2022). Positive/negative MPRA scores directly indicate increased/decreased regulatory activation due to sequence variants. In neuronal stem cells, SNPs with significant MPRA scores have a plateau distribution of Δ*repression* scores, unlike insignificant-MPRA-score SNPs (Figure 7A). More precisely, 52.4% and 42.3% of significant-MPRA-score enhancer and silencer SNPs were labeled as a raSNP, significantly higher than the 12.8% of all insignificant-MPRA-score SNPs (*p* < 10^−10^) and the 18.9% of insignificant-MPRA-score within enhancers and silencers (*p* < 10^−5^, Figure 7B).

**Figure 7.**
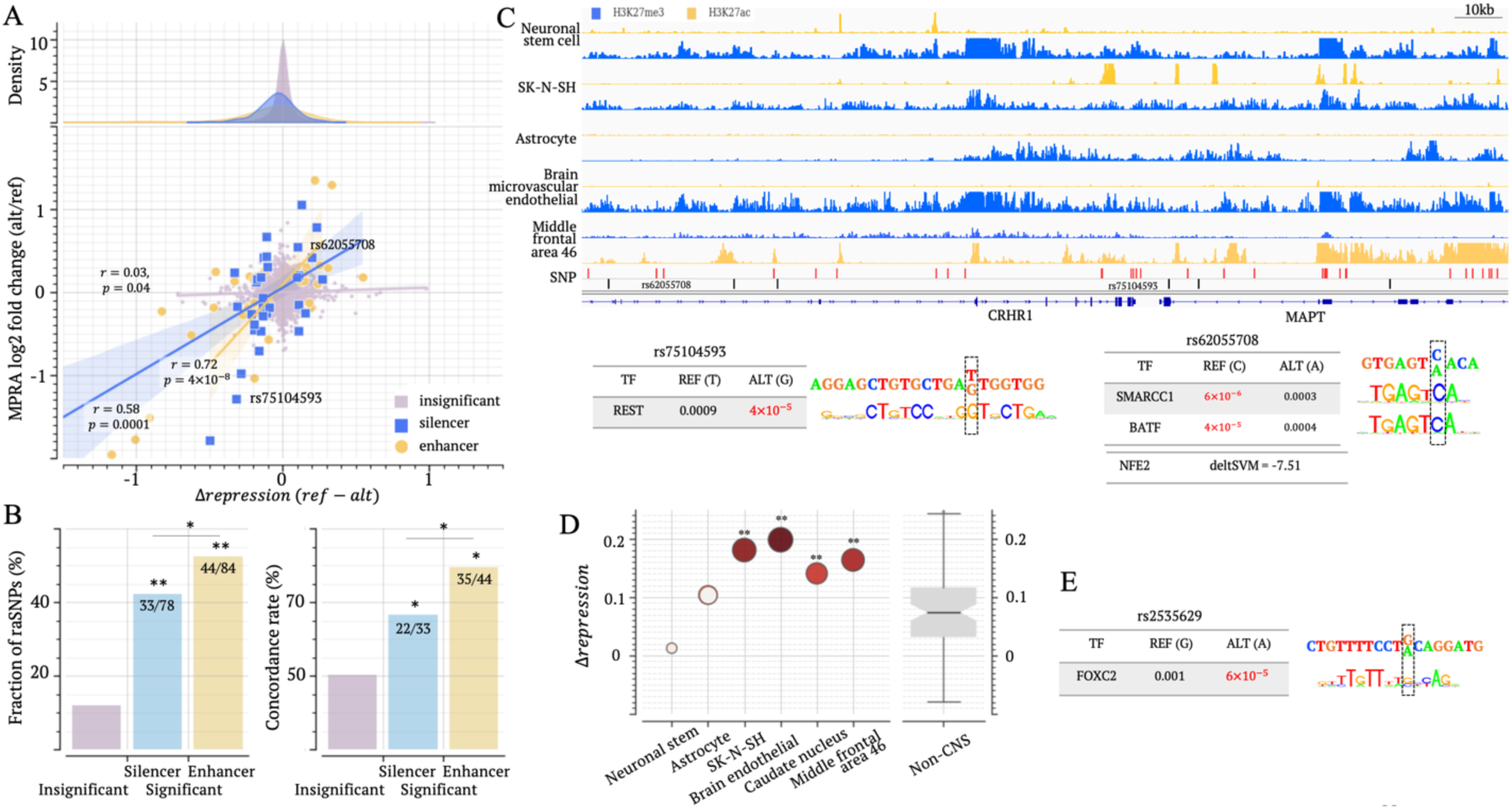
CNS raSNPs have strong regulatory impact. (A) Correlation between Δ*repression* and dementia MPRA scores in neuronal stem cell. Top panel illustrates Δ*repression* score distributions for different SNP groups. Bottom panel plot Δ*repression* and MPRA scores of SNPs. SNP groups here are insignificant-MPRA SNPs, significant-MPRA silencer, and enhancer SNPs. The analysis results in other CNS biosamples are presented in Figure S10. (B) Fractions of raSNPs (the left panel) and directional concordance between Δ*repression* and MPRA scores (the right panel) across SNP groups as defined in panel (A). The numbers of all examined SNPs, raSNPs, and concordant raSNPs are listed in the bars accordingly. (C) Epigenetic profile of silencer SNPs associated with PD in MAPT locus. TF binding motif mapping results on example SNPs are also presented. In the track of “SNP”, black and red bars represent tag PD SNPs and their LD SNPs (r^2^ > 0.8). (D) Δ*repression* scores of SCZ-associated SNP rs2533629 in CNS biosamples. (E) Analysis of TF binding motif mapping at rs2533629. ∗: *p* < 0.05 and ∗∗: *p* < 10^−5^.

Remarkably, Δ*repression* scores in neuronal stem cells positively correlate to MPRA scores. This positive correlation remains significant regardless of MRPA scores and SNP locations (*p* = 0.04, *r* = 0.03 among insignificant-MRPA-score SNPs; *p* = 0.0001, *r* = 0.58 among significant-MPRA-score silencer and *p* = 4 × 10^−8^, *r* = 0.72 among significant-MPRA-score enhancer SNPs, Figure 7A).

Among significant-MPRA-score silencer SNPs, Δ*repression* scores are directionally concordant to the corresponding MPRA scores in over two-thirds of instances (Figure 7B). This concordance rate is significantly higher than the 50% for insignificant-MPRA-score SNPs (binomial test *p* = 0.04). The robust correlation between Δ*repression* scores and MPRA scores is also evident in other CNS biosamples. The concordance rate is 67.5% among raSNPs (*p* = 10^−9^ vs 51.0% of insignificant-MPRA-score SNPs, Figure S21). Altogether, these findings strongly support the high accuracy of Δ*repression* scores in gauging the regulatory effects of variants, at least in CNS biosamples.

Focusing on specific SNPs, we started with the SNP rs62055708, which is associated with PD and many other neurological traits, including autism, bipolar disorder, brain volume measurement, and intelligence. It’s a SNP located within candidate silencers in most CNS biosamples except the middle frontal area (Figure 7C). The C to A change at this SNP has Δ*repression* = 0.20 in neuronal stem cells, aligning with an MPRA-score of 0.42. Also, this SNP corresponds to reduced significance in binding motif mapping for transcriptional repressors SMARCC1 (the allele C vs A: *p* = 6 × 10^−6^ vs 0.0003) and BATF (*p* = 4 × 10^−5^ vs 0.0004, Figure 7C, see Methods)(Schaniel et al. 2009; Li et al. 2012). Additionally, as predicted by SNP-SELEX deltaSVM (Yan et al. 2021), the change from the allele C to A at this SNP gains a binding site for NFE2, a transcriptional activator as discussed above (Figure 6). Another PD-associated SNP is rs75104593. Consistent MPRA-score = –1.28 and Δ*repression* = −0.32 in neuronal stem cells suggest that the substitution at this SNP (from T to G) boosts the repressive effect, which could be supported by the increased significance of binding motif mapping for REST, a well-known repressor TF (Figure 7C). It is worth noting that both REST and NFE2 are widely recognized as PD-associated factors (Bento-Pereira and Dinkova-Kostova 2021; Brent et al. 2021), further strengthening the connection between these two raSNPs and PD.

At a SCZ-associated rs2535629, a substitution from G to A has been experimentally confirmed to increase the binding affinity of CTCF in a ChIP-Allele-Specific-qPCR assay (ChIP-AS-qPCR) and diminish the suppressive impact in a dual-luciferase reporter gene assay (Li et al. 2022). This SNP is a raSNP located within candidate silencers in four out of six examined CNS biosamples. The Δ*repression* scores in CNS biosamples are significantly higher than in non-CNS biosamples (Student’s *t*-test *p* = 10^−21^, Figure 7D). TF-motif-mapping analysis also shows increased binding affinity of FOXC2 due to the G to A change at this SNP (Figure 7E). FOXC2 is a transcription activator contributing to gene overexpression in various cancers, like glioblastoma (Li et al. 2013). This finding provides an additional mechanistic clue to understanding the potential role of rs2535629 in the development of SCZ. The strong agreement of Δ*repression* with MPRA scores and TF binding affinity prediction underscores the high accuracy of Δ*repression* scores in assessing the regulatory impact of genetic variants.

### T1D and other autoimmune diseases are linked to variants in candidate silencers

To assess Δ*repression* scores in immune biosamples, we compared them with MPRA scores measured in lymphoblastoid cell lines from two independent studies, i.e., the multiplex MPRAs, denoted as mMPRA below (Tewhey et al. 2016) and the variant-based MPRAs, referred to as vMPRA (Abell et al. 2022).

SNPs with significant mMPRA scores show a higher magnitude of Δ*repression* than insignificant-mMPRA-score SNPs (Figures 8A and S22). Specifically, 37% and 36% of significant-mMPRA-score SNPs in candidate silencer and enhancer are raSNPs in immune biosamples, significantly surpassing the 19% of insignificant-mMPRA-score SNPs (*p* < 10^−10^, Figure 8B). Notably, Δ*repression* scores in immune cells are significantly positively correlated with mMPRA scores across different SNP sets (*p* < 10^−10^ across insignificant-mMPRA-score and candidate silencer/enhancer significant-mMPRA-score SNPs).

**Figure 8.**
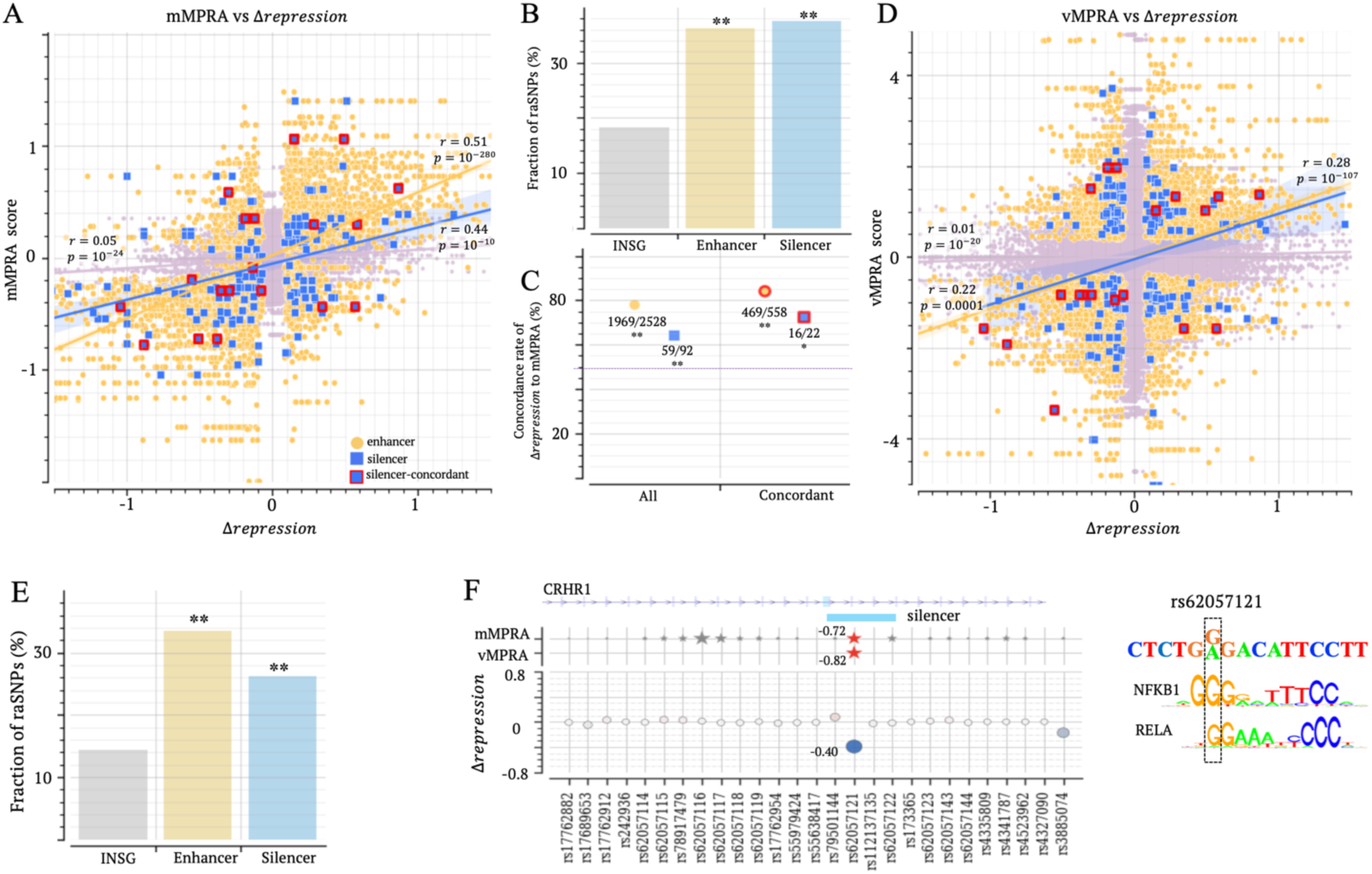
Immune raSNPs within candidate silencers have strong regulatory impact. (A) Correlations between Δ*repression* and mMPRA scores in SNP groups. Silencer-concordant represents the SNPs where significant mMPRA and vMPRA scores directionally align. (B) Fractions of raSNPs among insignificant-mMPRA, significant-mMPRA silencer and significant-mMPRA enhancer SNPs. (C) Concordance rate between Δ*repression* and mMPRA score across SNP groups. “All” represents all significant-mMPRA SNPs in candidate silencers or enhancers. “Concordant” is as denoted in (A). Numbers alongside each marker indicate the count of SNPs showing concordant Δ*repression* and mMPRA scores, as well as the total number of SNPs considered. The dashed line represents the expectation when randomly shuffling Δ*repression* scores. (D) Correlations between Δ*repression* and vMPRA scores. (E) Fractions of raSNPs among insignificant-vMPRA, significant-vMPRA silencer and significant-vMPRA enhancer SNPs. (F) Δ*repression*, mMPRA, vMPRA scores on the T1D-associated rs62057121 and its neighboring SNPs. In the top panel, red/grey stars indicate significant/insignificant mMPRA or vMPRA scores, respectively. All significant scores are listed next to the corresponding markers. In addition, the TF binding motif analysis on this SNP is presented. ∗: *p* < 0.05 and ∗∗: *p* < 10^−8^

Furthermore, 72.3% of raSNPs in candidate silencers have a Δ*repression* score directionally concordant to their mMPRA scores, significantly exceeding the 49.4% as expected from randomly shuffling Δ*repression* scores, as well as the 51.4% of SNPs with insignificant MPRA scores (*p* < 0.01, Figure 8C). This concordance rate further increases to 78.9% among the SNPs where mMPRA and vMPRA scores directionally align, although these increases are not significant most likely due to the shrinking size of the analyzed SNP set (Figure 8C). Similar trends are mirrored among enhancer SNPs. Additionally, Δ*repression* scores exhibit significant positive correlations with vMPRAs in immune biosamples (*p* < 0.0005, Figures 8D and S23). For example, 33% and 26% of significant-vMPRA-score SNPs in candidate silencers and enhancers are raSNPs in immune biosamples, significantly surpassing the 14% of insignificant-vMPRA-score SNPs (*p* < 10^10^, Figure 8E). These significant correlations and high concordance rates are in the line with the observations on dementia MPRAs (Figure 7), generalizing the high validity of Δ*repression* scores in evaluating regulatory effects of variants across different biosample groups.

For example, rs6207121, a SNP associated with T1D, exhibits significant scores in mMPRA and vMPRA. This SNP, with Δ*repression* = −0.51, is detected as a raSNP within a candidate silencer in CD4+ alpha-beta T cells, holding the highest magnitude within its 4kb vicinity. This Δ*repression* score directionally aligns with the corresponding mMPRA and vMPRA scores (Figure 8F). Moreover, the analysis of binding motif mappings suggests that this variant potentially disrupts a binding site for NFKB1, a key TF known for dual repressive and activating functions in the immune system (Li and Verma 2002) and in the development of T1D (Konrad et al. 2019).

Another example is the rs242561 SNP, which has been linked to a range of immune and neurological disorders, including T1D, BPD, and Parkinson’s disease. This SNP is predicted as a raSNP in both immune and CNS biosamples. The significantly negative Δ*repression* scores in CNS biosamples correlate with the negative dementia MPRA score (Figure S24). Interestingly, this SNP is located within a DFRE, acting as a silencer in immune biosamples but an enhancer in CNS biosamples, likely by recruiting different TFs in immune cells and in neurons.

## Discussion

Here, we report 2.8 million candidate silencers in 97 human biosamples representing diverse origins, collectively spanning 19.4% of the human genome. More than half of candidate silencers (55%) are DFRE, acting as enhancers in alternative biosamples, which evidences the widespread presence of DFREs. Furthermore, the majority (67%) of hESC candidate silencers function as DFREs, which could still increase after additional human biosamples are explored. In differentiated cells, the hESC-silencer-converted enhancers exhibit a notable enrichment in TFBSs of CTCF, RAD21 and ZNF143, as well as in chromatin contacts, suggesting they frequently act as anchors for chromatin contacts.

This study demonstrates the vital role of candidate silencers in complex diseases with a strong genetic basis. This new perspective goes beyond GWAS, uncovering how individual disease-associated genes are regulated during pathogenesis. For example, SCZ and BPD have been linked through GWAS to the dysregulation of neuronal differentiation and apoptosis. Our analysis shows that this dysregulation may primarily stem from variants within CNS candidate silencers. Moreover, the disruption of the GABAergic interneuron has been reported as a key cause in SCZ (Nakazawa et al. 2012). Our analysis further underpins that the variants within CNS candidate silencers could be responsible for this disruption. Similarly, in the gene loci of *INS* and *IGF2*, T1D-associated SNPs are greatly concentrated within candidate silencers, implying the pivotal roles that candidate silencers play in regulating these genes in the immune system. Silencer variants thereby greatly account for the dysregulation of these two genes in the context of T1D (Steck and Rewers 2011). Collectively, silencers represent fundamental components underlying the development of many complex diseases. The profiles of silencers (along with enhancers) can facilitate the unraveling of genetic basis of these diseases.

It is important to note that this study is centered around silencers, with enhancers serving as a reference point. The goal is to underscore the significance of silencers in disease research, rather than to provide an exhaustive genetic portrait of diseases. Genetic components of diseases that go beyond these elements are not within the scope of this study. For example, we do not delve into *LILR* genes, which host TAK-associated variants in their promoters (Yoshifuji and Terao 2020). Evidently, a comprehensive understanding of a polygenetic disease requires the exploration of diverse regulatory elements, along with protein-coding variants, which is the motivation of this study.

We further extended the analysis to the level of individual genetic variants. High correlations with the experimental results from MPRA and SNP-SELEX studies validate the accuracy of Δ*repression* scores in predicting the regulatory impact of SNPs across different biosamples. RaSNPs, the SNPs having a significant Δ*repression* score, frequently hold significant MPRA scores and SNP-SELEX scores, confirming the substantial impact of these variants on disease susceptibility. Prioritizing disease-causal SNPs is the initial step to reveal molecular mechanisms underlining polygenetic diseases. Delineating the cascading effects of these SNPs, such as how they alter TF binding affinity, chromatin organization and gene expression, represents the subsequent challenge. It is noteworthy that, although we present experimental and computational results of TF binding affinities of raSNPs here, this issue will remain incompletely addressed until experimental profiling of TF binding expands to many more TFs and spans additional cell types across multiple developmental time points. For example, as demonstrated here, experimental results from SNP-SELEX assays are restricted to a small proportion of SNPs, possibly due to their cell specificity (Yan et al. 2021).

Here, silencer identification primarily relies on H3K27me3 ChIP-seq peaks. While this histone mark is a well-characterized and widely-accepted proxy of repressive regulatory influence, our candidate silencer profiles might be incomplete due to the existence of non-H3K27me3 silencers (Doni Jayavelu et al. 2020; Pang and Snyder 2020). The strong association of candidate silencers with developmental genes, particularly those active during embryonic stages, aligns with the established role of H3K27me3 in developmental processes (Ngan et al. 2020). This association may also hint at a possible bias toward H3K27me3 among candidate silencers. Currently, the detection of non-H3K27me3 silencers are limited to few cell types (Pang and Snyder 2020; Hussain et al. 2023; Xiusheng et al. 2023) and/or confined to certain genomic regions (Grass et al. 2003; Mouri et al. 2023), which largely hampers the investigation on these silencers. Despite these constraints, our analysis underscores the significance of silencers in controlling key biological processes and highlights their profound influence in disease susceptibility.

## Methods

### Identification of candidate silencers

We trained the TREDNet model, a two-phase deep learning model (Hudaiberdiev et al. 2023) to predict enhancers and silencers. We downloaded DNase-seq peaks, H3K27ac and H3K27me3 ChIP-seq peaks (“narrow peak”) for 111 biosamples from ENCODE project (https://www.encodeproject. org/, Table S1). Enhancer training sequences were defined as the DNase-seq peaks overlapping H3K27ac ChIP-seq peaks but not H3K27me3 peaks in the central 400bp. Silencer training sequences were defined as the DNase-seq peaks overlapping H3K27me3 peaks but not H3K27ac peaks in the central 400bp as well as the H3K27me3 peaks not overlapping H3K27ac peaks. To accommodate this multi-label classification task, the output layer of TREDNet models consist of three nodes with the activation function of “softmax”, representing silencer, enhancer, and control samples, respectively. The cost function used here is “categorical cross entropy”. We held out chromosomes 7 and 8 for testing. All other autosomes were used for building the classification model (Hudaiberdiev et al. 2023). Consequently, testing sequences, having no overlap with training sequences, provide an unbiased computational evaluation on the performance of the TREDnet models.

For silencer prediction, 1kb-long input sequences were evaluated by silencer prediction scores. The cutoff for labeling silencers (say *t*_s_) was set as a false positive rate (FPR) of 0.1 in test samples, with control samples to candidate silencers in the ratio of 9:1. DNase-seq peaks or H3K27me3 ChIP-seq peaks that have a silencer score greater than *t*_s_ were predicted as silencers. Similarly, the cutoff for labeling enhancers (say *t*_e_) was set as a false positive rate (FPR) of 0.1 in test samples, again with control samples to candidate enhancers in the ratio of 9:1. DNase-seq peaks that have an enhancer score greater than *t*_e_were predicted as enhancer. The sequences marked as both enhancers and silencers were considered as “uncertain”, which account for less than 1% of silencers or enhancers in all tested biosamples and were excluded from further analysis. To this end, 97 biosamples have over-5000 candidate enhancers and over-5000 candidate silencers, which were investigated in this study.

Each candidate enhancer/silencer is 1kbp-long. A candidate silencer in a biosample was considered as a DFRE if it overlaps with an enhancer in another biosample by over-200 bp. Similarly, an enhancer was considered as a DFRE when it overlaps with a candidate silencer in another biosample by over-200 bp.

### GWAS SNP enrichment in individual gene loci

We assess the significance of GWAS SNPs associated with a disease (*i*) in a gene locus (*j*), *p_ij_*, in comparison to the whole genome using the binomial test. The gene loci having a *p_ij_*. < 10^−8^ are regarded as associated with the disease *i*. Similarly, in a disease-associated locus (say *j*), the enrichment of given GWAS SNPs within silencers, 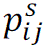, is assessed by using the binomial test. That is,

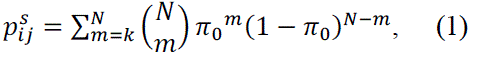

where π_0_is the ratio of the locus length to the whole genome. *N* and *k* are the total number of given GWAS SNPs within the candidate silencers and the number of given GWAS SNPs within the candidate silencers in the locus *j*. The enrichment of given GWAS SNPs in candidate enhancer in the locus *j* is evaluated by replacing *N* and *k* in Eq. (1) with the number of given GWAS SNPs within the enhancers in the whole genome and in the locus *j*, respectively.

### Δ*repression*

To evaluate the regulatory impact of a variant with the wild type (wt) and mutant allele (mu), we input the 1kb-long sequences centering at this variant to a trained TREDNet model. We then obtained the silencer and enhancer prediction scores for all alleles. The false positive rates of silencer prediction scores (denoted as *FPR*^+^) are evaluated based on test samples with the size ratio of control samples to candidate silencers of 9:1. Similarly, the false positive rates corresponding to enhancer prediction scores (represented by *FPR^e^*), are evaluated based on test samples. The regulatory alteration between these alleles is then estimated as

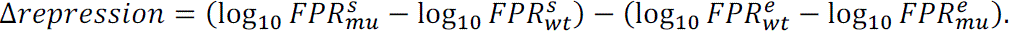

A positive Δ*repression* indicates a decrease in the repressive impact due to the mutation.

In a biosample, we evaluated the significance *p* value of a Δ*repression* score by comparing with Δ*repression* scores on all common SNPs documented in dbSNP as of 2017 (Sherry et al. 2001). A Δ*repression* score is regarded as significant if *p* < 0.05 among all common SNPs. A SNP is marked as raSNP if the corresponding Δ*repression* score is significant. When analyzing the correlation between between Δ*repression* and MPRA scores (Figures 7 and 8), SNPs are considered as a silencer SNP either when they are located within a candidate silencer or when they overlap with a H3K27me3 ChIP-seq peak and have 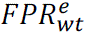 < 0.05. Similarly, SNPs are considered as an enhancer SNP either when they are located within a candidate enhancer or when they overlap with a H3K27ac ChIP-seq peak and have 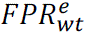 < 0.05.

### Data and tools

We downloaded GWAS SNPs curated in the National Human Genome Research Institute (NHGRI) catalog (McMahon et al. 2018) and in UK Biobank release 2 cohort (Bycroft et al. 2018). All the GWAS SNPs associated with the same trait, according to their Experimental Factor Ontology ID (Malone et al. 2010), were merged into one SNP set. We extended trait-associated SNP sets by including the SNPs in tight linkage disequilibrium (LD r^2^ > 0.8) to GWAS SNPs based on EUR population in 1000 Genomes Project. To this end, we retrieved a total of 2.2 million GWAS SNPs, which are associated with 2,212 distinct traits. Among these traits, 1,166 traits are linked to more than 80 SNPs and thus used in our investigation.

Hi-C chromatin contacts were downloaded from the study by Salameh et al. (Salameh et al. 2020). Brain volume measurements include intracranial, hippocampal, thalamus and subiculum volume measurement. The sets of GWAS SNPs associated with these traits significantly overlap among each other (Jaccard similarity > 0.65), and therefore were merged as brain-volume-associated SNPs in this study.

We evaluated the correlations between Δ*repression* and SNP-SELEX scores for each TF in each tested biosample. In a biosample, TFs having at least 10 SNPs holding significant SNP-SELEX and significant Δ*repression* scores were included to ensure a robust estimation on the correlation between Δ*repression* and SNP-SELEX scores.

TF ChIP-seq data used here were downloaded from the ENCODE project (Table S2). TF binding motif were downloaded from the MEME Suite (https://meme-suite.org/meme/db/motifs). Find Individual Motif Occurrence (FIMO), with the default setting, was used to find the mappings of binding motifs in given sequences (Grant et al. 2011).

## Declarations

### Funding

This research was supported by the Intramural Research Programs of the National Library of Medicine, National Institutes of Health

## Data Availability

All data produced in the present work are contained in the manuscript.

## Acknowledgements

This work utilized the computational resources of the NIH HPC Biowulf cluster (http://hpc.nih.gov).

## Authors’ contributions

D.H. performed computational analysis and analyzed the data. I.O. supervised computational work. D.H. prepared figures and tables. D.H. and I.O. wrote the manuscript.

## Competing interests

The authors declare that they have no competing interests.

### Availability of data and material

Please see the section of “Data and Tools” and supplementary materials.

## Ethics approval and consent to participate

Not applicable.

## Consent for publication

Not applicable.

## Supplementary Notes

### Gene expression profiles

We obtained the gene expression data from the ENCODE project ^1^ for 215 biosamples (Table S4). We used gene annotations from the GENCODE ^2^ to define the transcription start site for each gene. A candidate silencer or enhancer was associated with its nearest gene. Gene expression levels were normalized as the fold change to the average expressions across biosample. That is, for a gene (say *i*) and its expression level in a biosample *b* (say, *e_i,b_*), its normalized expression level *ne_i,b_* was calculated as 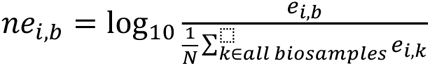, N = the number of biosamples.

### Locus-specific enrichment of silencers and enhancers

We used gene annotations from GENCODE ^2^. The locus of a gene encompasses the gene body along with its two flanking upstream and downstream intergenic regions. Using this annotation, there are 26,550 distinct gene loci in the human genome. For a given gene locus (say, g) and a biosample, the count of candidate silencers located within this locus was tallied, and the silencer enrichment significance was determined using the binomial test, i.e.,

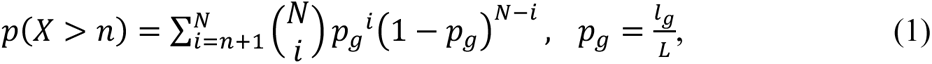

where n and N are the numbers of silencers within the gene locus g and in the whole genome, respectively. *l_g_* and L denote the length of the locus g and the whole genome, respectively. Using Bonferroni multiple-testing correction, the silencer density in the locus g is regarded as significantly higher than expected in the whole genome when *p*(*X* > *n*) ≤ 0.05/*G*. Here, G is the total number of gene loci in the whole genome. Similarly, the significancy of enhancer enrichment in a gene locus is assessed based on enhancer counts.

### eQTLs

We downloaded eQTL data from the GTEx project ^3^ for 17 distinct tissues, comprising 13 brain tissues, colon, lung, spleen, and whole blood. For each GTEx tissue, we checked the distribution of eQTLs within candidate silencers in the corresponding biosamples. For example, we gathered eQTLs from all brain GTEx tissues and examined their density within candidate silencers in each brain biosample. In the end, 40 biosamples were tested in this analysis.

## Supplementary Figures

**Figure S1.**
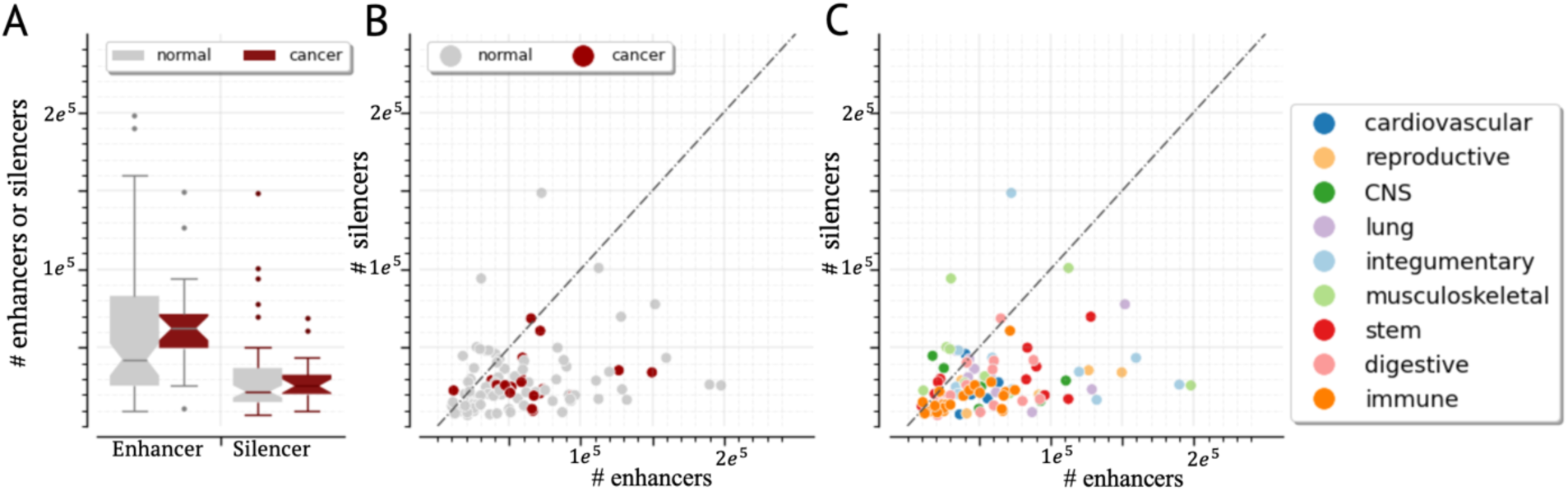
Numbers of candidate silencers and enhancers across biosamples. Each dot represents a biosample.

**Figure S2.**
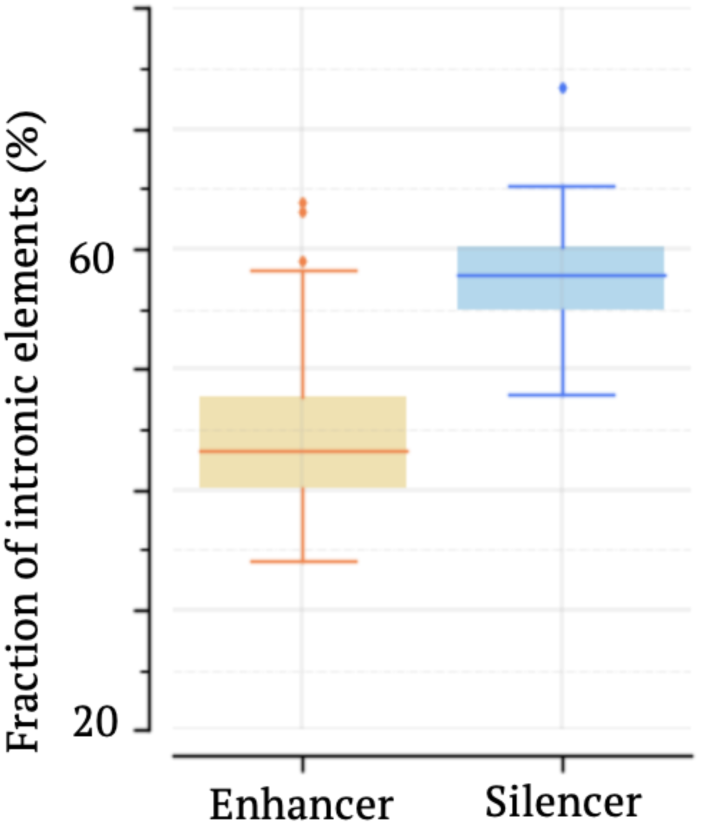
Fractions of intronic candidate silencers and enhancers.

**Figure S3.**
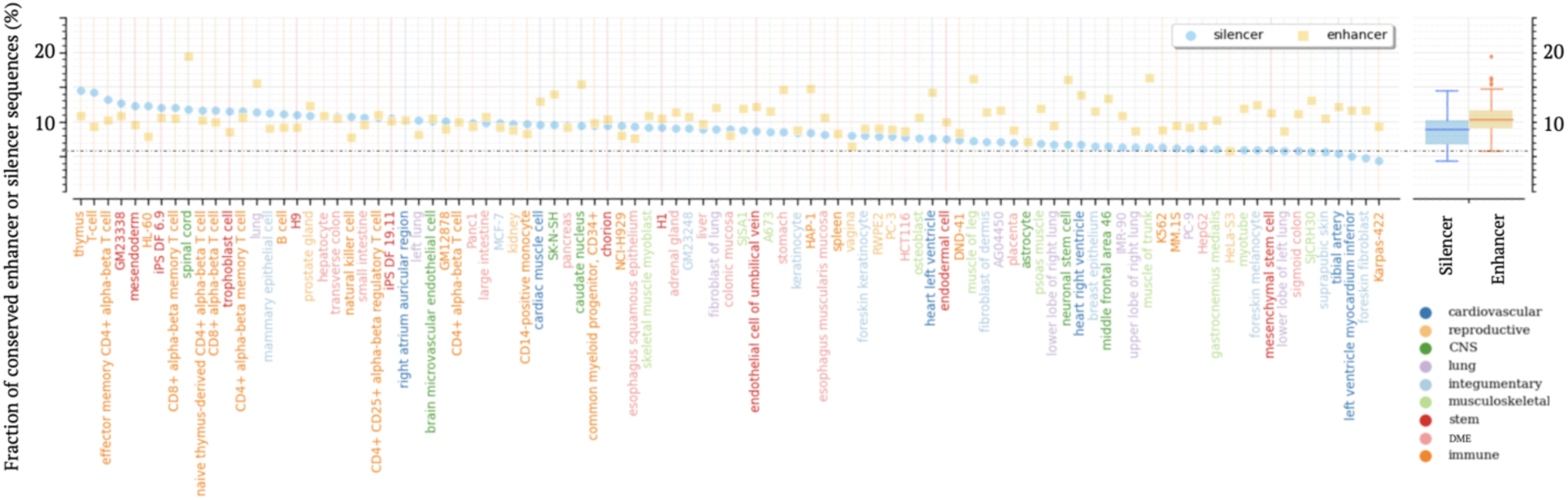
Fractions of conserved candidate silencers or enhancers. DME represents “digestive and metabolic and endocrine” biosample categories.

**Figure S4.**
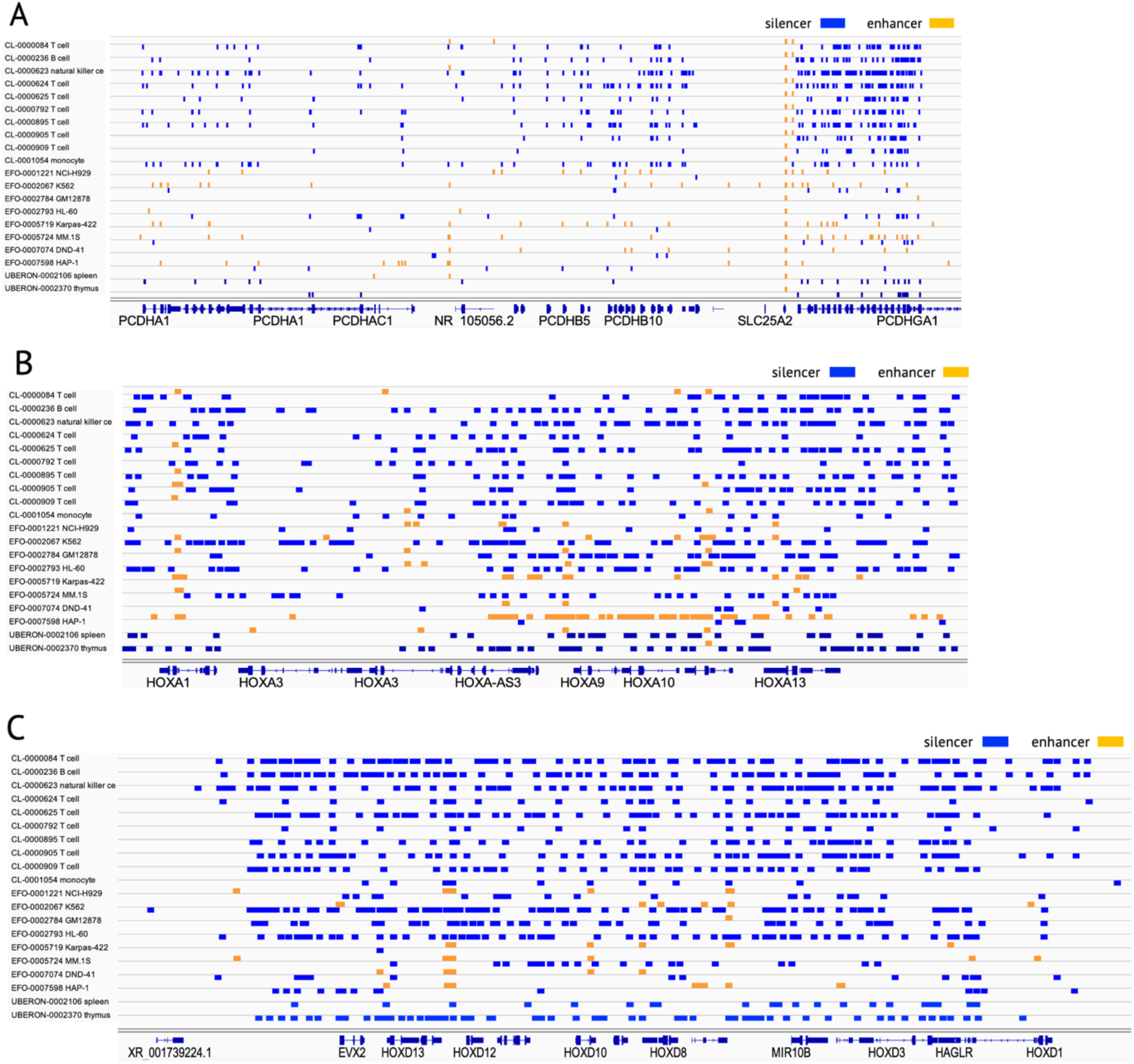
Profiles of candidate silencer and enhancer across immune biosamples in. (A) the loci of *PCDHA/B* genes, (B) the loci of *HOXA* genes, (C) the loci of *HOXD* genes. All these gene loci are enriched with candidate silencers in immune biosamples.

**Figure S5.**
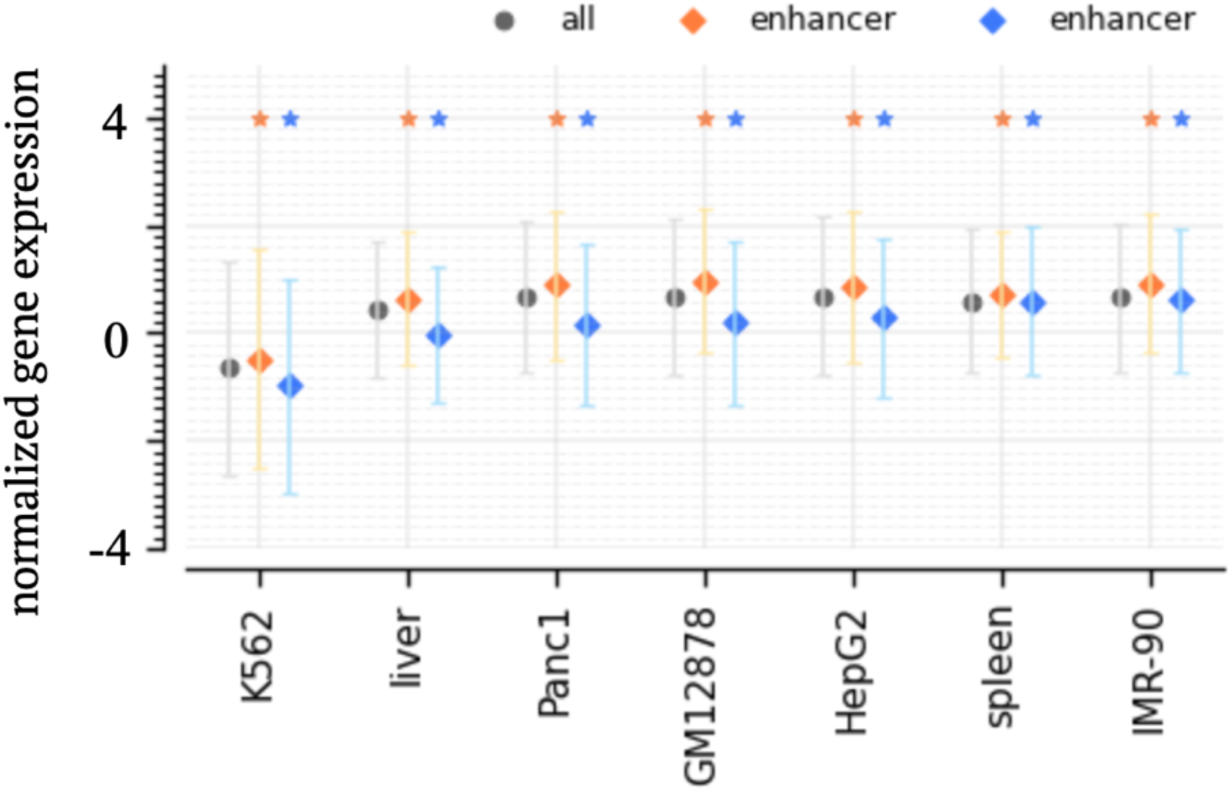
Expression of genes in contact with candidate silencers. The markers and their flanking lines represent the medians and standard deviations of gene expression levels. Blue and orange asterisks denote significantly low and high expression levels, respectively, compared to those of all assayed genes (p<0.05).

**Figure S6.**
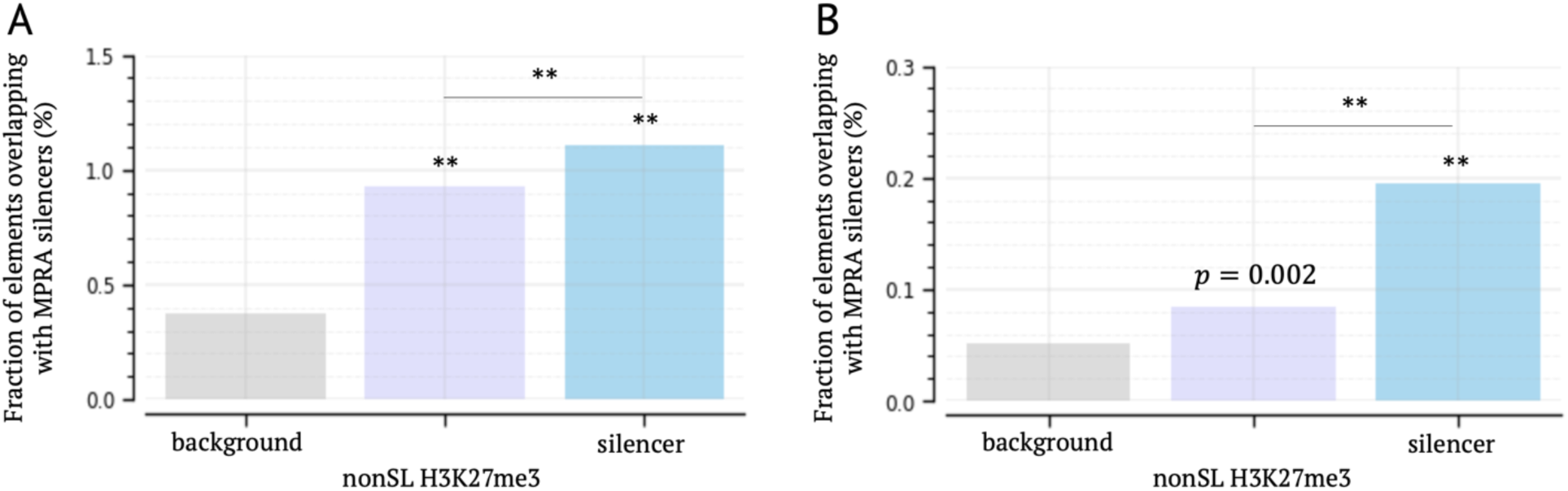
Significant overlap between candidate silencers and MPRA experimentally validated silencers in the biosamples. (A) K562 and (B) HepG2. ∗∗: *p* < 10^*$%^. “nonSL H3K27me3” represents the H3K27me3 ChIP-seq peaks not overlapping with candidate silencers.

**Figure S7.**
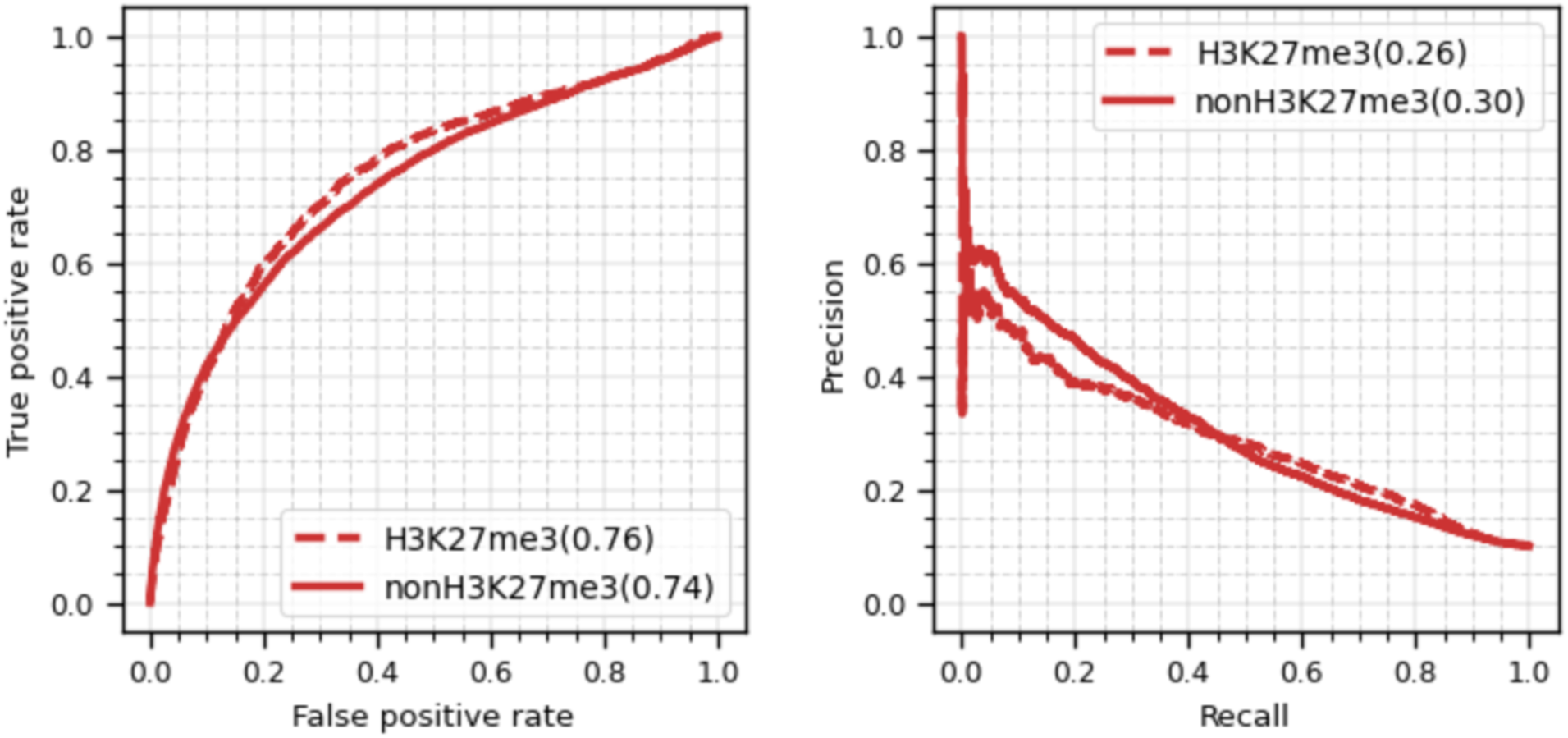
Classification performance of the TREDnet model on MPRA silencers with and without H3K27me3 ChIP-seq peaks.

**Figure S8.**
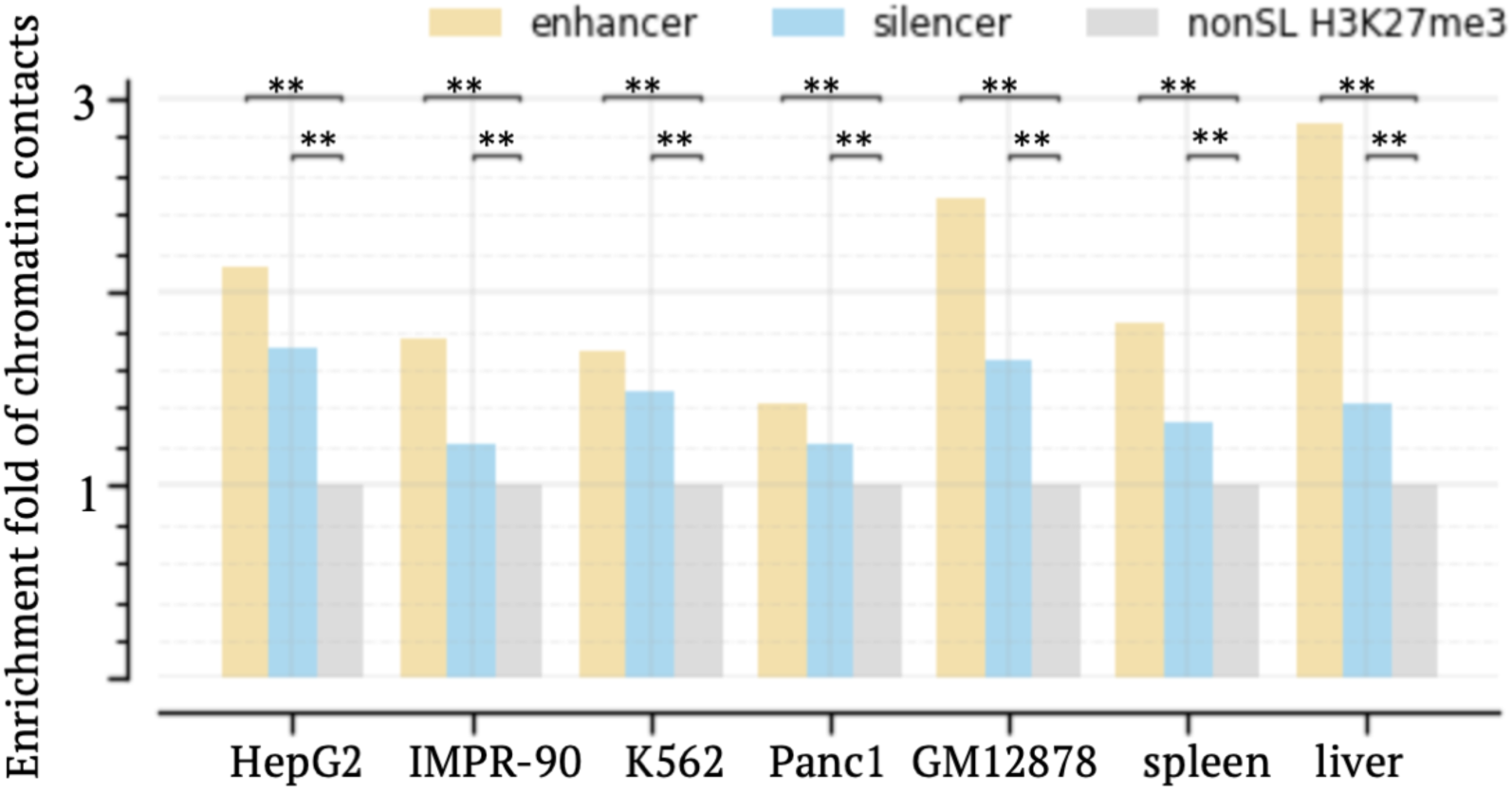
Enrichment of chromatin contacts in candidate silencers, enhancers and H3K27me3 ChIP-seq peaks not overlapping with candidate silencers (represented as nonSL H3K27me3). ∗∗: *p* < 10^*$%^.

**Figure S9.**
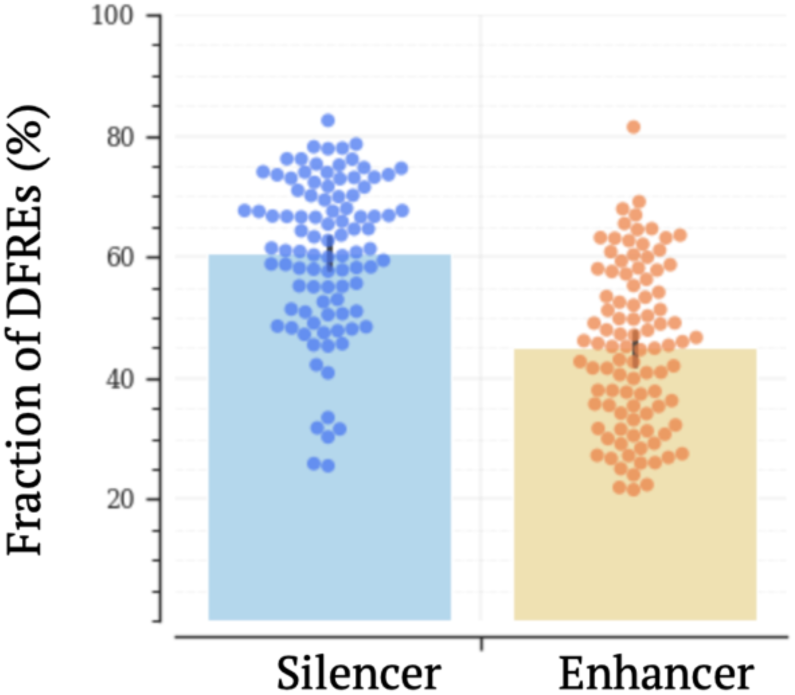
Fractions of DFREs among candidate silencers and enhancers across biosamples.

**Figure S10.**
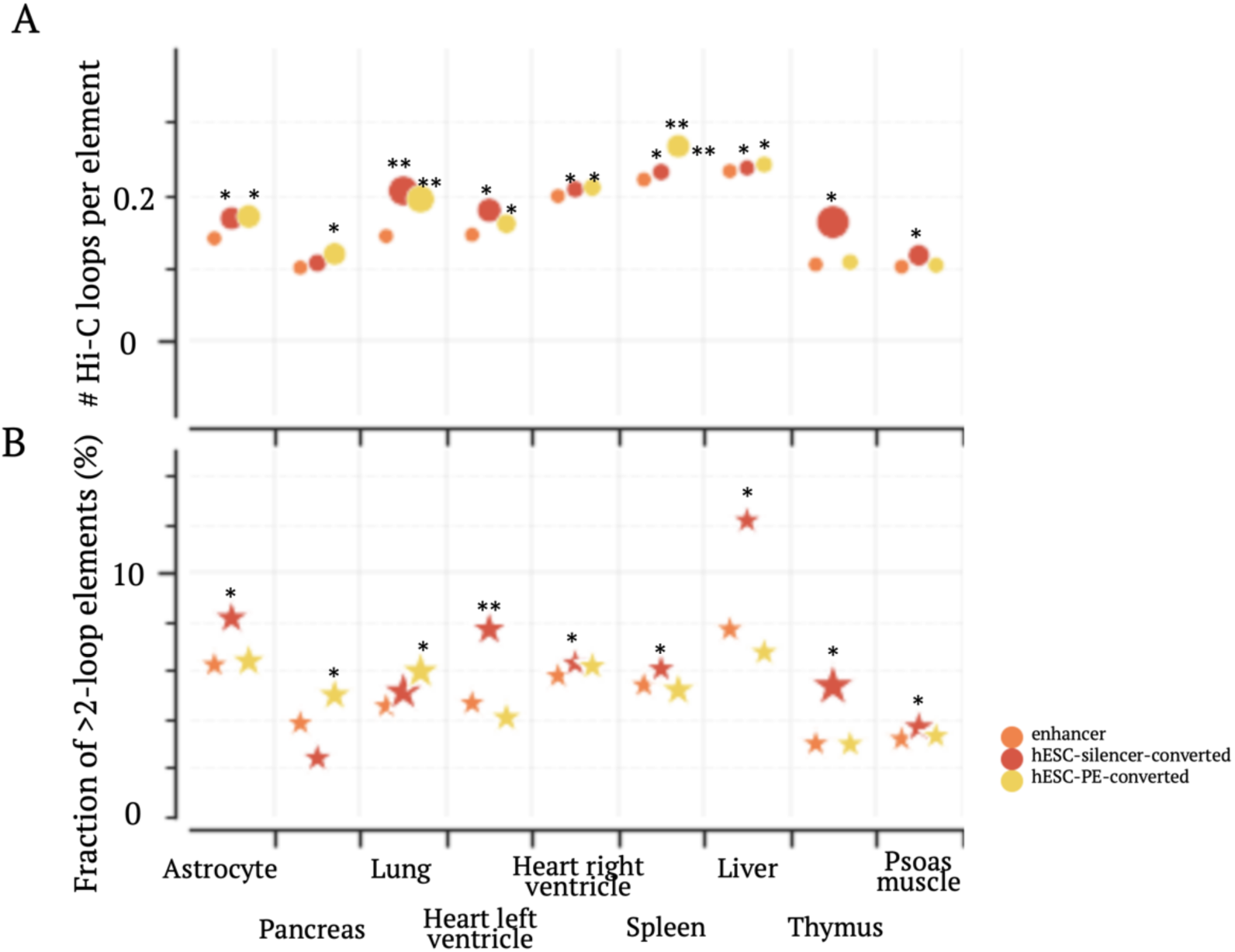
Density of chromatin contacts across enhancer groups. (A) Numbers of chromatin contacts. (B) Fractions of elements having >2 contacts. ∗ *p* < 10^−3^ and ∗∗ *p* < 10^−8^.

**Figure S11.**
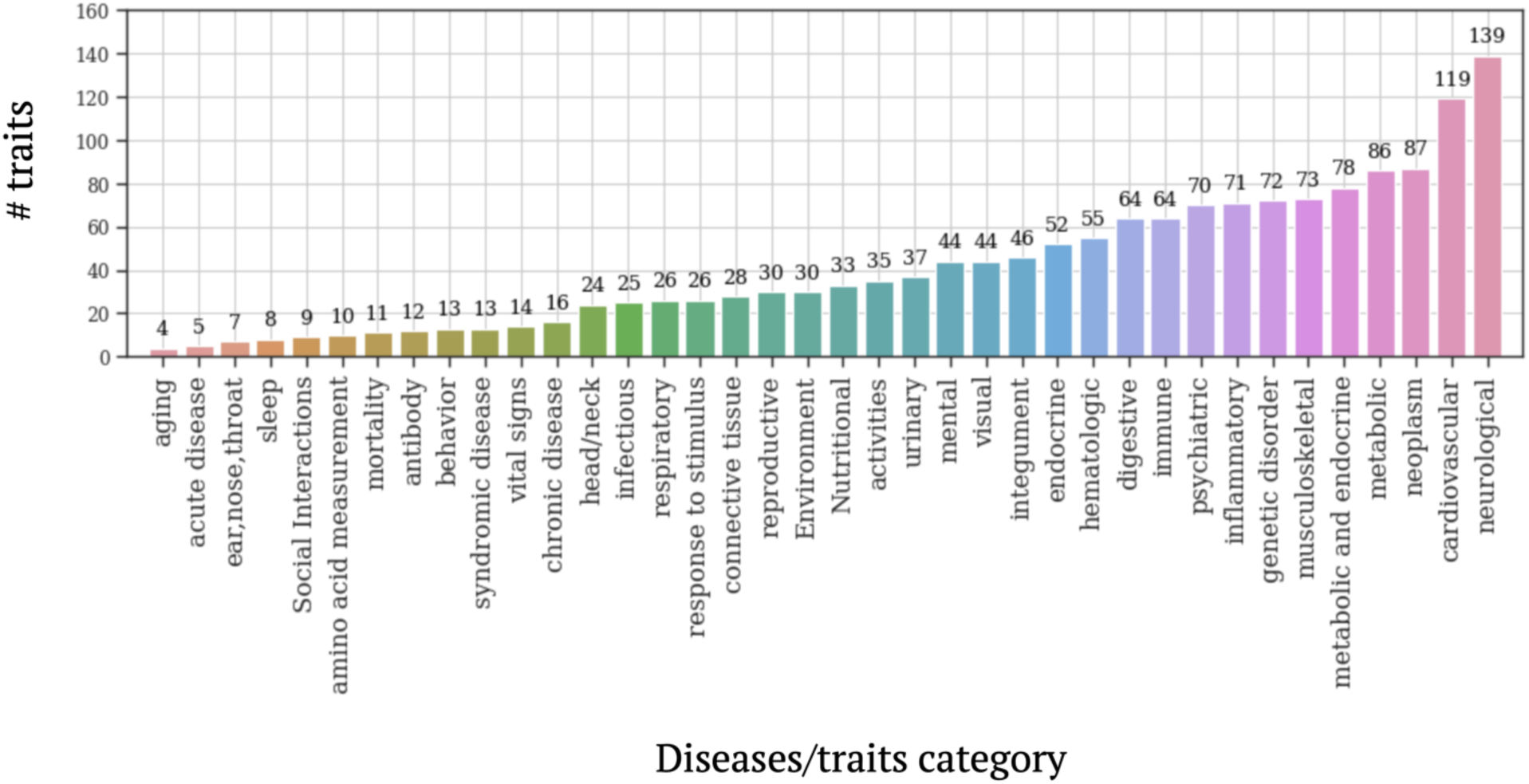
The distribution of GWAS traits investigated in this study.

**Figure S12.**
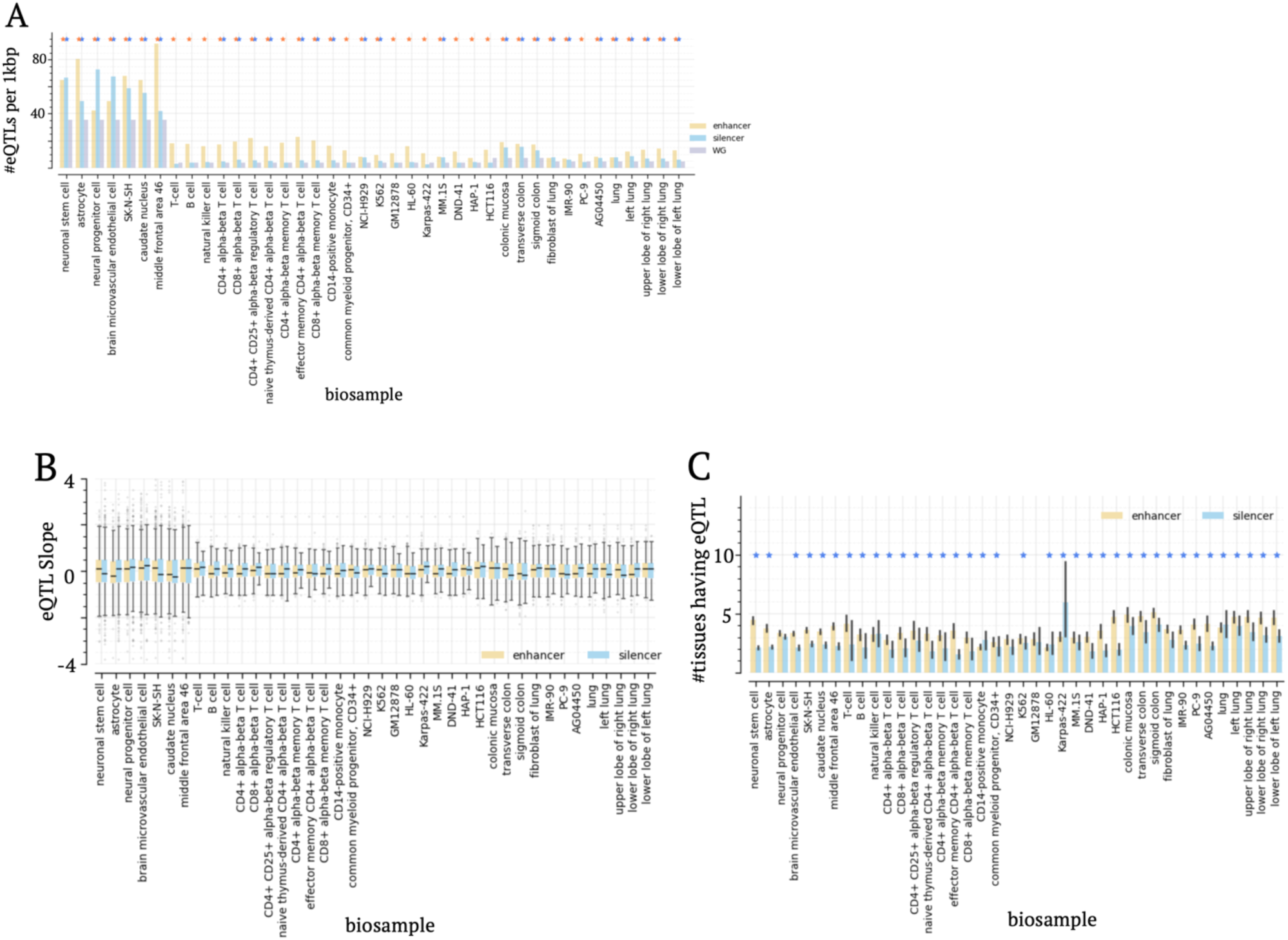
Distribution of eQTLs. (A) Densities, (B) eQTL slopes, and (C) tissue-specificities of eQTLs within candidate silencers and enhancers. The asterisks in (A) represent the significant enrichment compared to the whole genome (p<0.05). The asterisks in (C) represent the significant tissue-specificity levels of candidate silencer eQTLs compared to those of enhancer eQTLs. For a given eQTL, a low number of tissues in which this eQTL was detected suggests its high tissue specificity.

**Figure S13.**
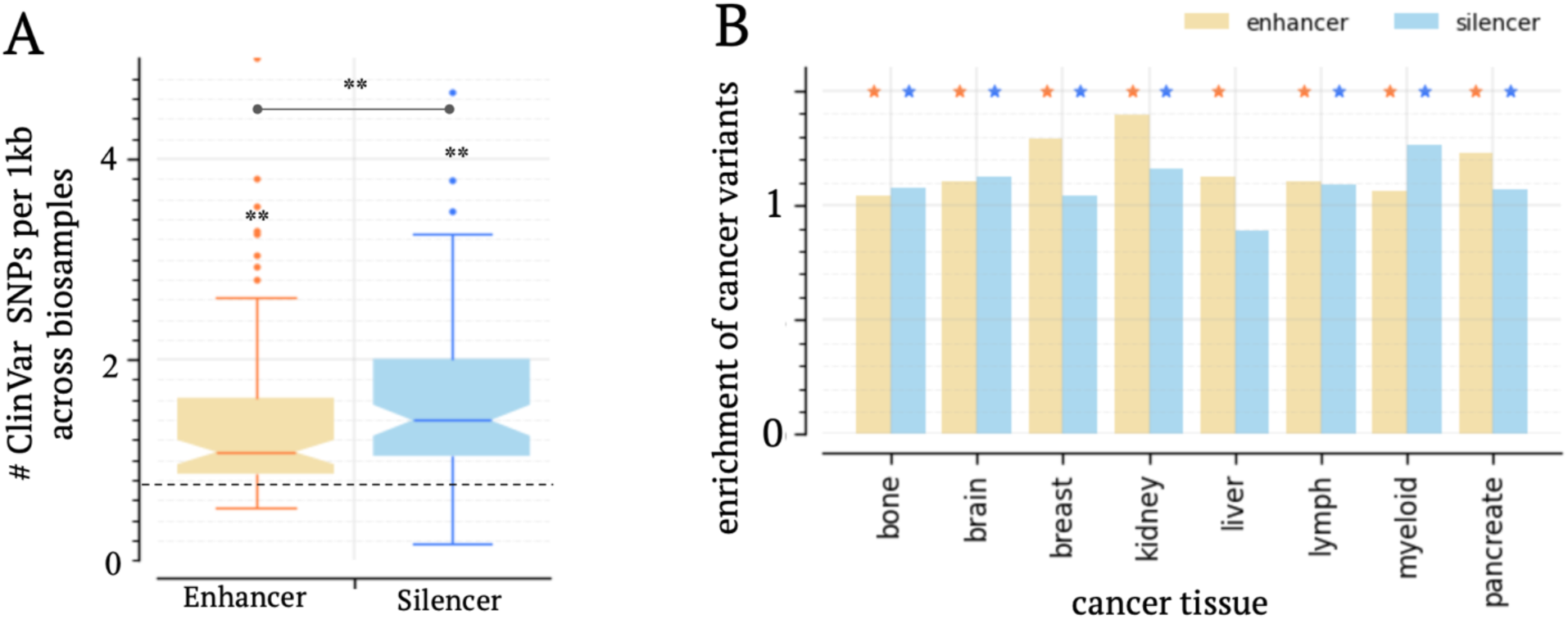
Enrichment of ClinVar SNPs and cancer somatic variants in candidate silencers. (A) Densities of ClinVar SNPs in candidate silencers and enhancers across all tested biosamples. The dash line represents the density of ClinVar SNPs in the whole genome. ∗∗ *p* < 10^*0^. (B) Densities of cancer somatic variants within candidate silencers in matched biosamples. The blue and orange asterisks represent a significant enrichment within candidate silencers and enhancers compared to the whole genome (p<0.05), respectively.

**Figure S14.**
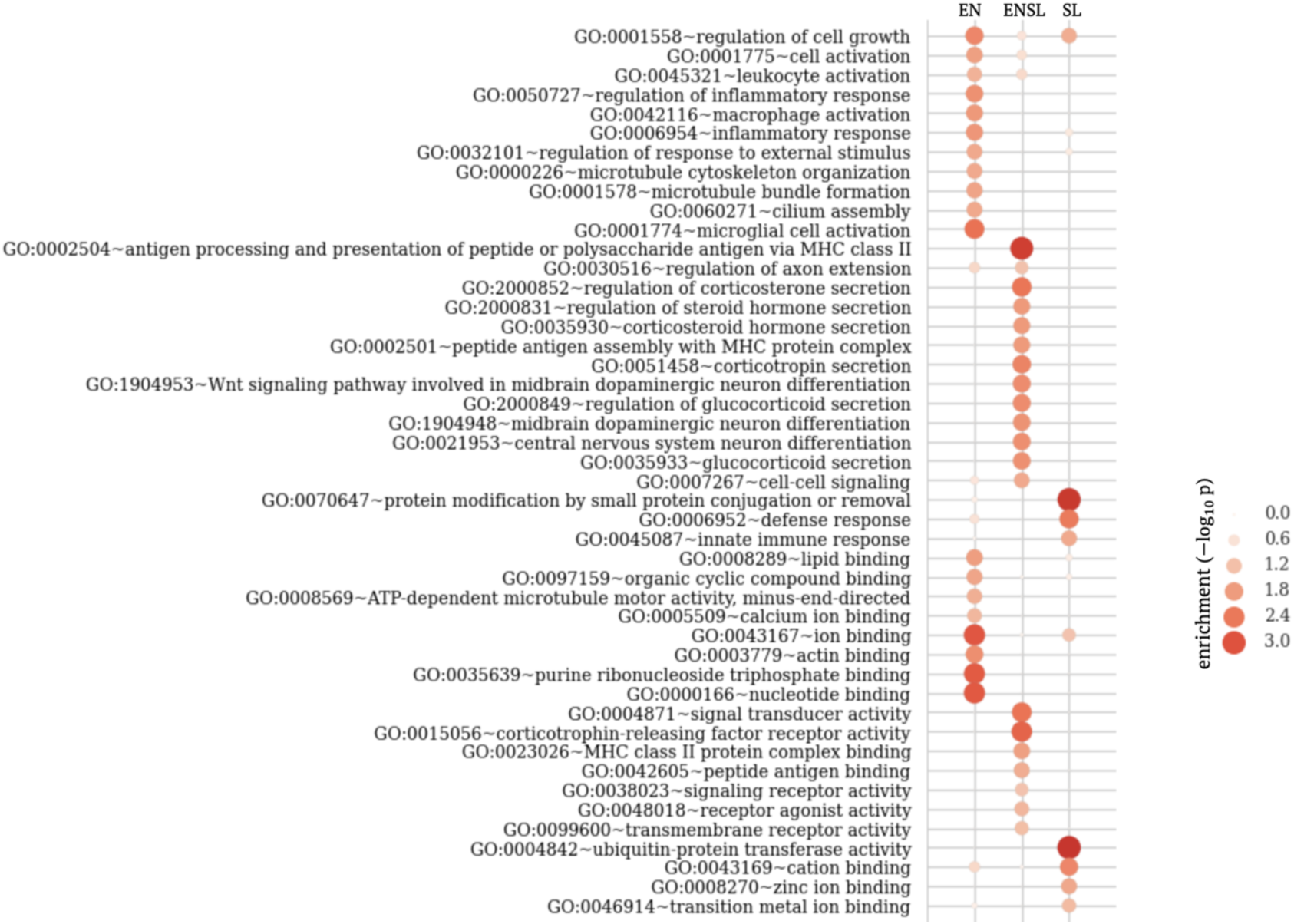
Function analysis of PD-associated gene groups defined in Figure 4C.

**Figure S15.**
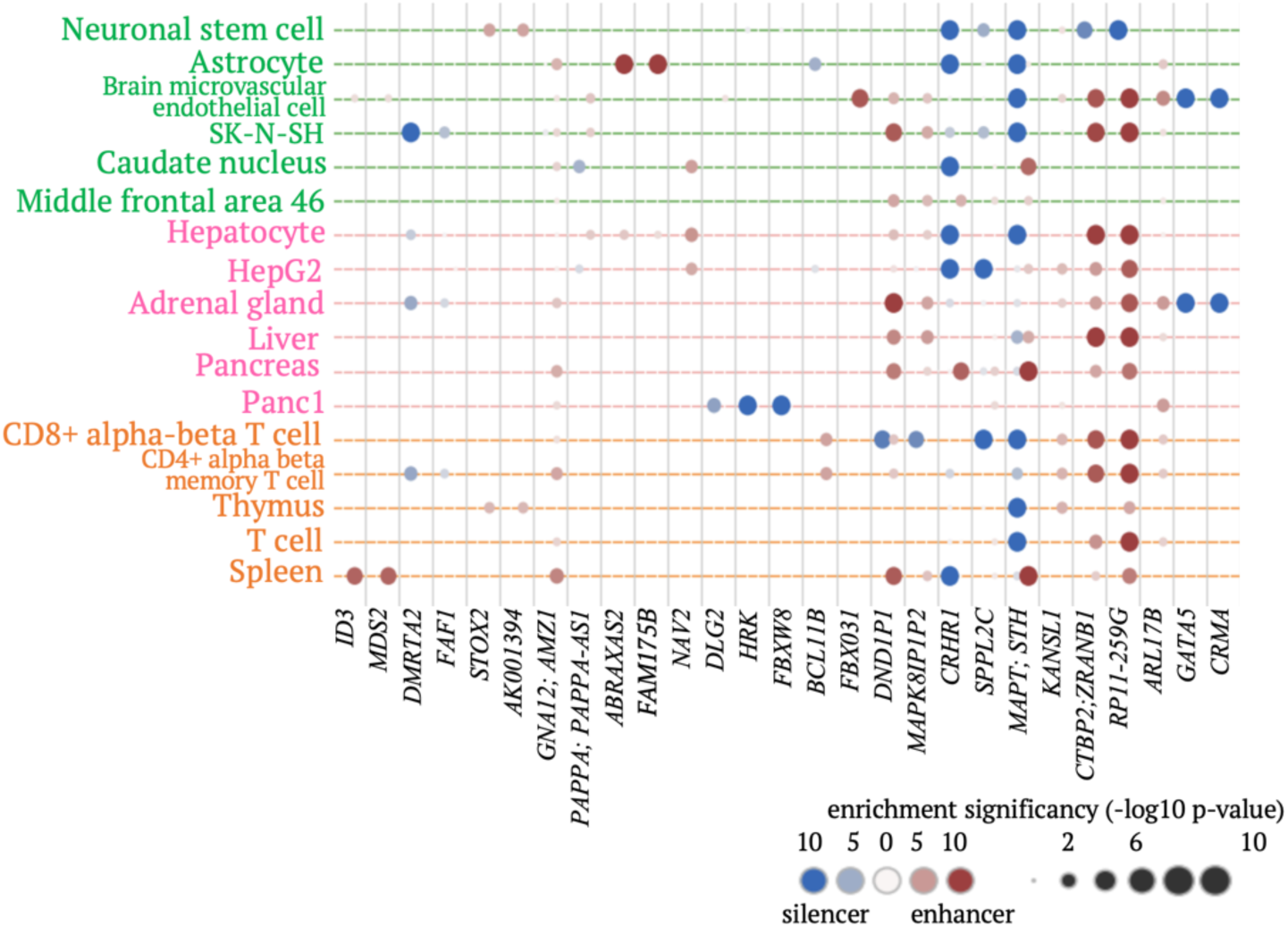
Enrichments of brain-volume-associated SNPs within candidate silencers and enhancers across gene loci. Gene loci having significant enrichment of the examined SNPs within either candidate silencers or enhancers are included in the plot.

**Figure S16.**
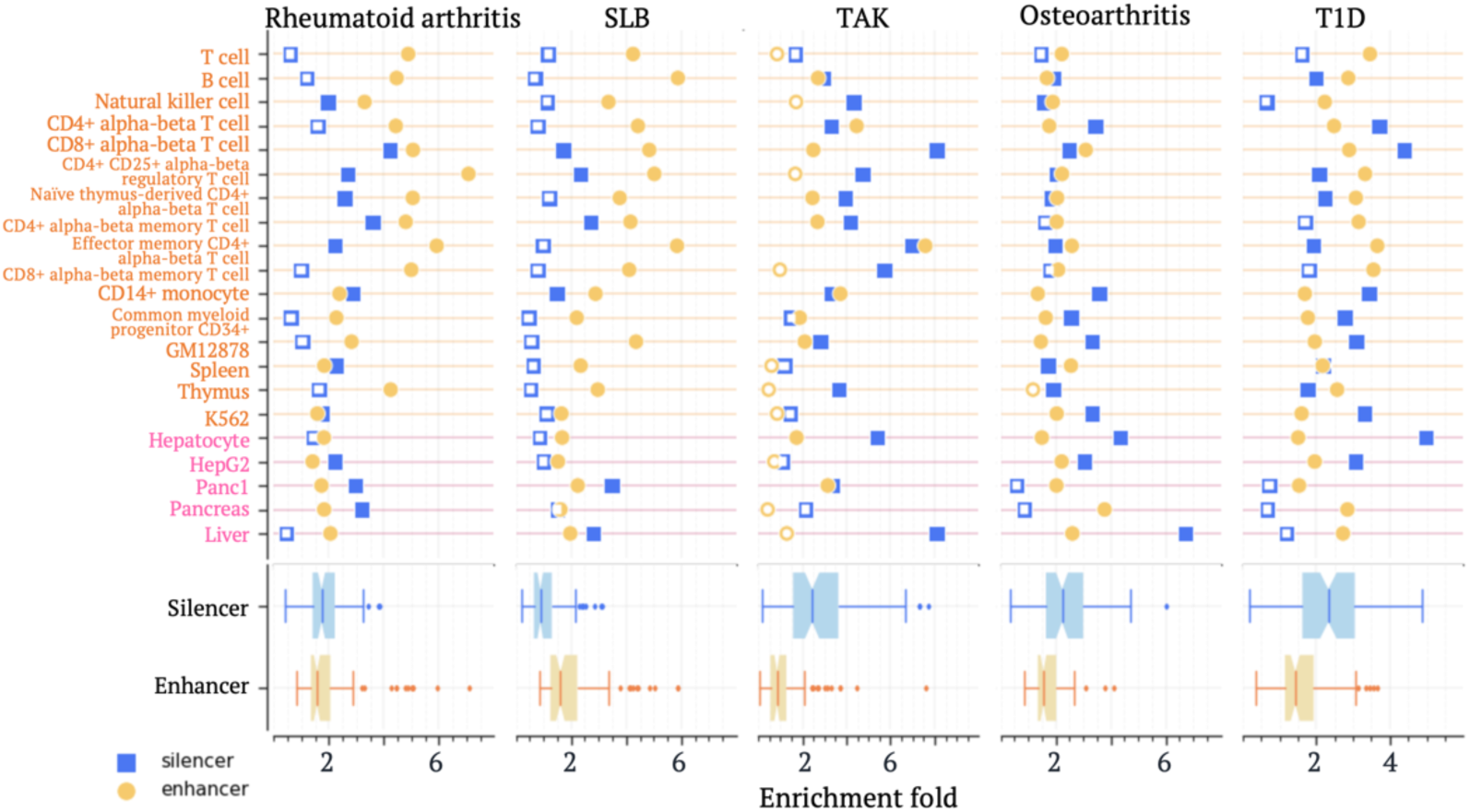
Enrichments of SNPs associated with autoimmune diseases. Enrichment folds are estimated in comparison to the whole genome. Significant enrichments are denoted by solid markers (*p* < 10^−5^).

**Figure S17.**
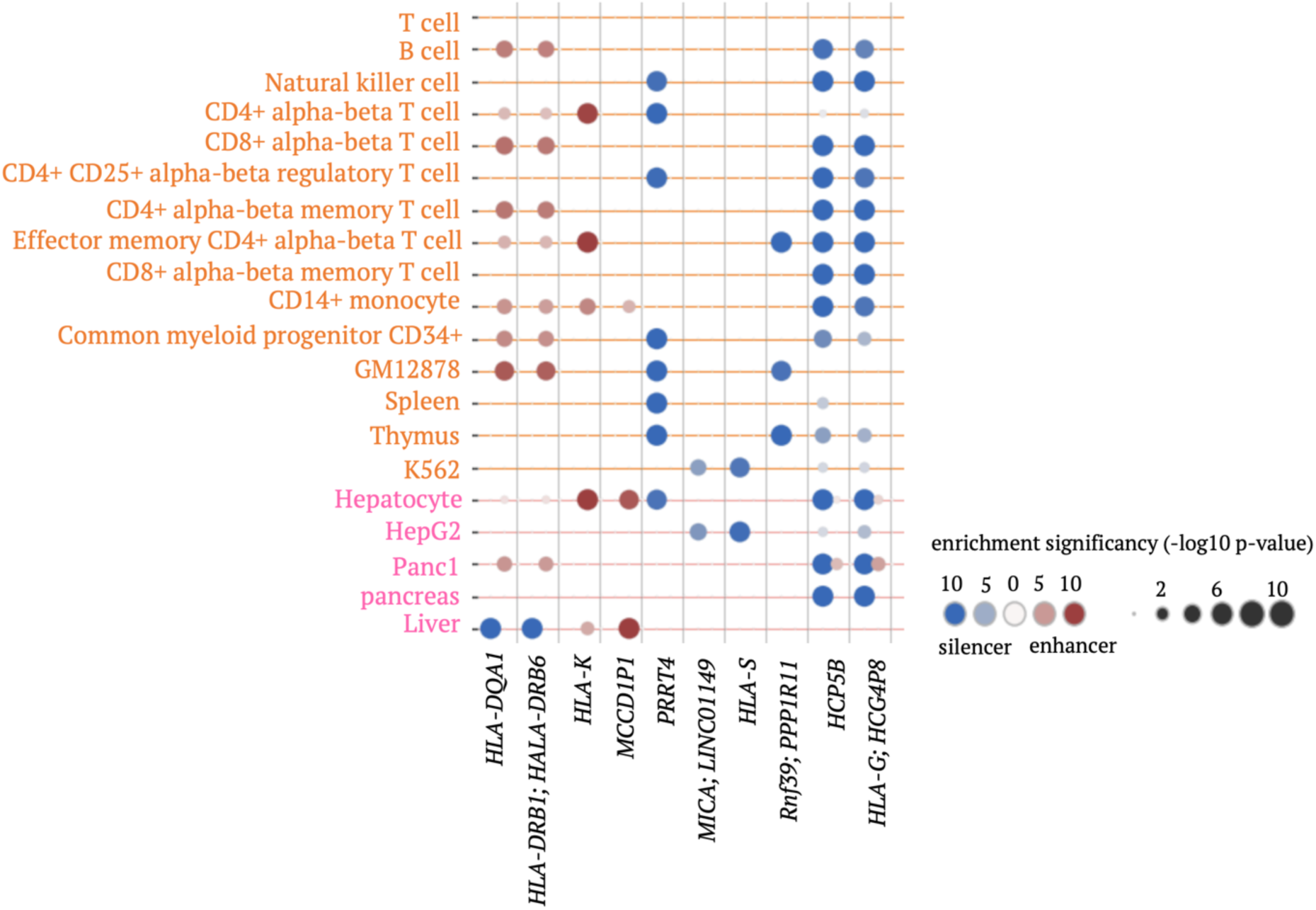
Enrichments of TAK-associated SNPs within candidate silencers and enhancers in gene loci. Gene loci having significant enrichment of the examined SNPs within either silencers or enhancers are included in the plot.

**Figure S18.**
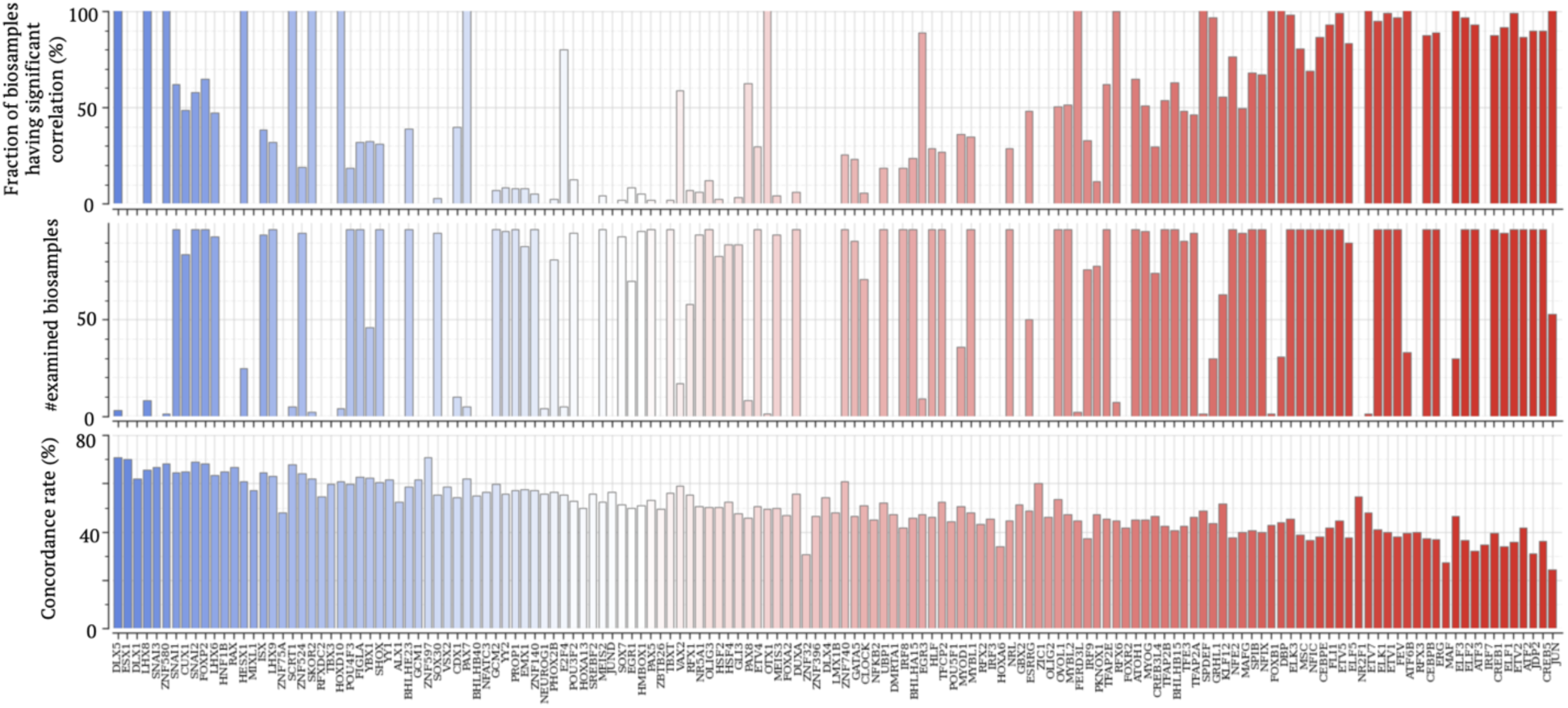
Distribution of significant correlations between Δ*repression* and SNP-SELEX scores for each TF across biosamples. For each TF, the top panel presents the number of biosamples for which SNP-SELEX scores of this TF significant correlate with Δ*repression*. The middle panel presents the number of biosamples for which this TF was examined. The bottom panel presents the concordance rate between Δ*repression* and SNP-SELEX scores.

**Figure S19.**
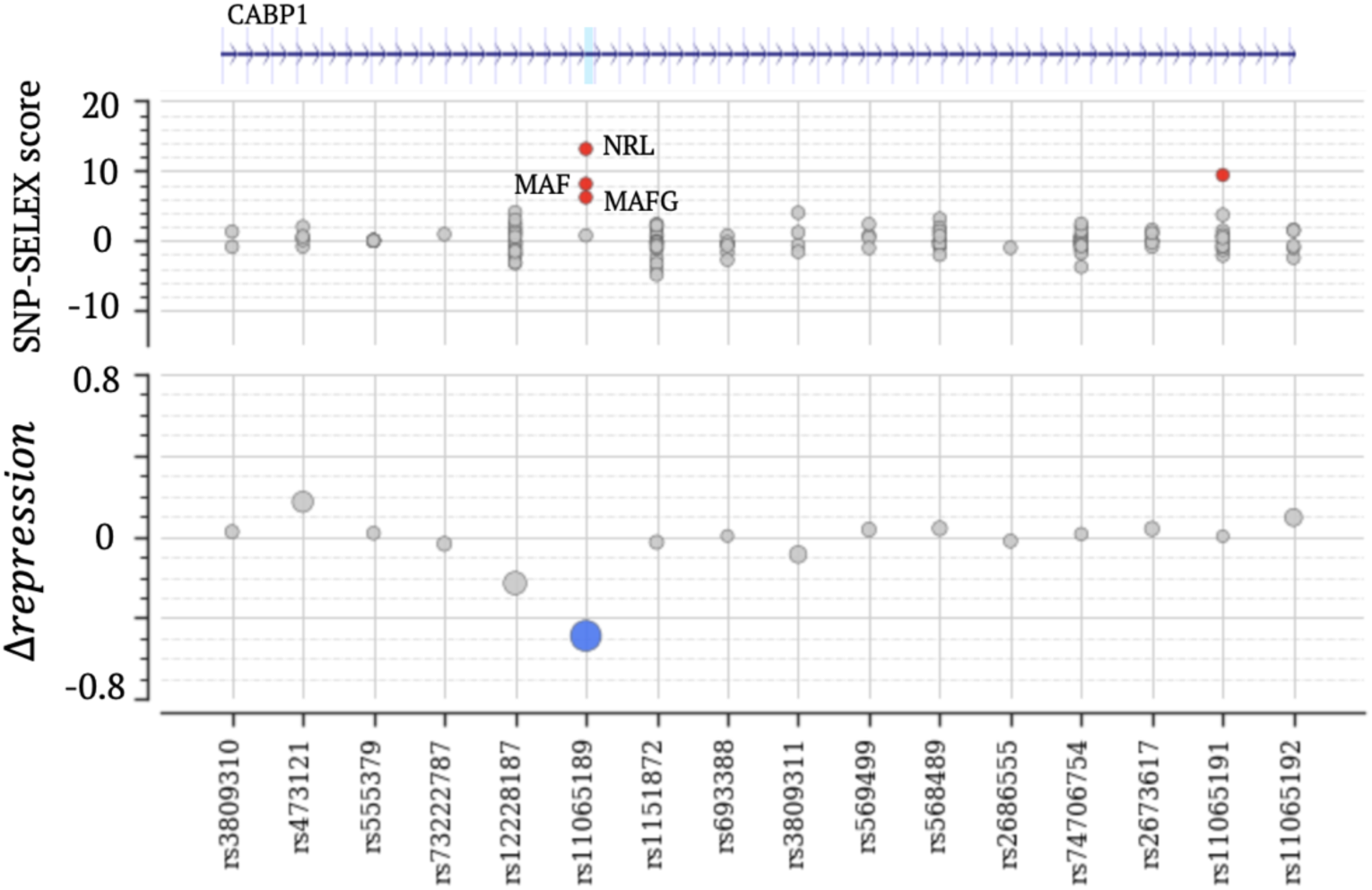
Δ*repression* and SNP-SELEX scores on the SCZ-associated rs11065189 and its neighboring SNPs. In the top panel, red/grey dots indicate significant/insignificant SNP-SELEX scores. TFs corresponding to the significant scores are listed. In the bottom panel, blue/grey dots indicate significant/insignificant Δ*repression* scores.

**Figure S20.**
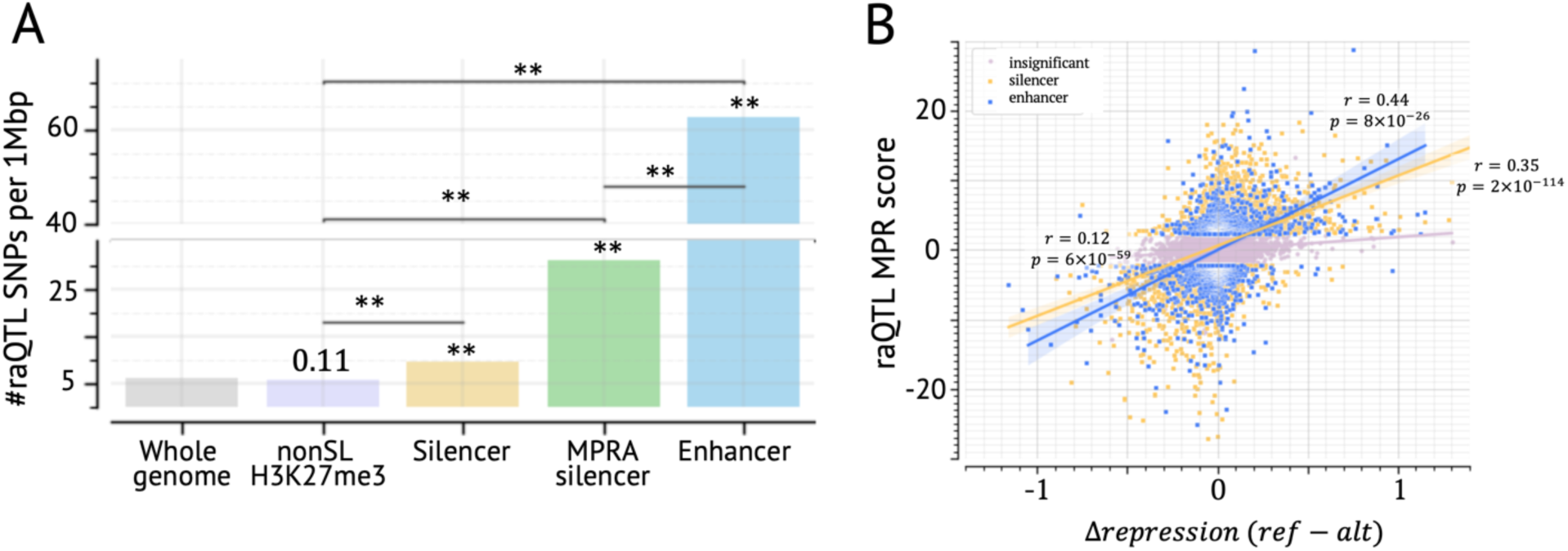
Analyses based on raQTLs. (A) raQTLs are enriched in the predicted silencers and MPRA silencers as compared to the expected across the whole genome and within non-predicted-silencer H3K27me3 ChIP-seq peaks (labelled as nonSL H3K27me3 here). Asterisks and the number over the bars suggest the enrichment p value as compared to the whole genome. ∗∗: *p* < 10^*$%^. (B) Δ*repression* scores significantly correlate with raQTL scores, regardless in predicted silencers or enhancers.

**Figure S21.**
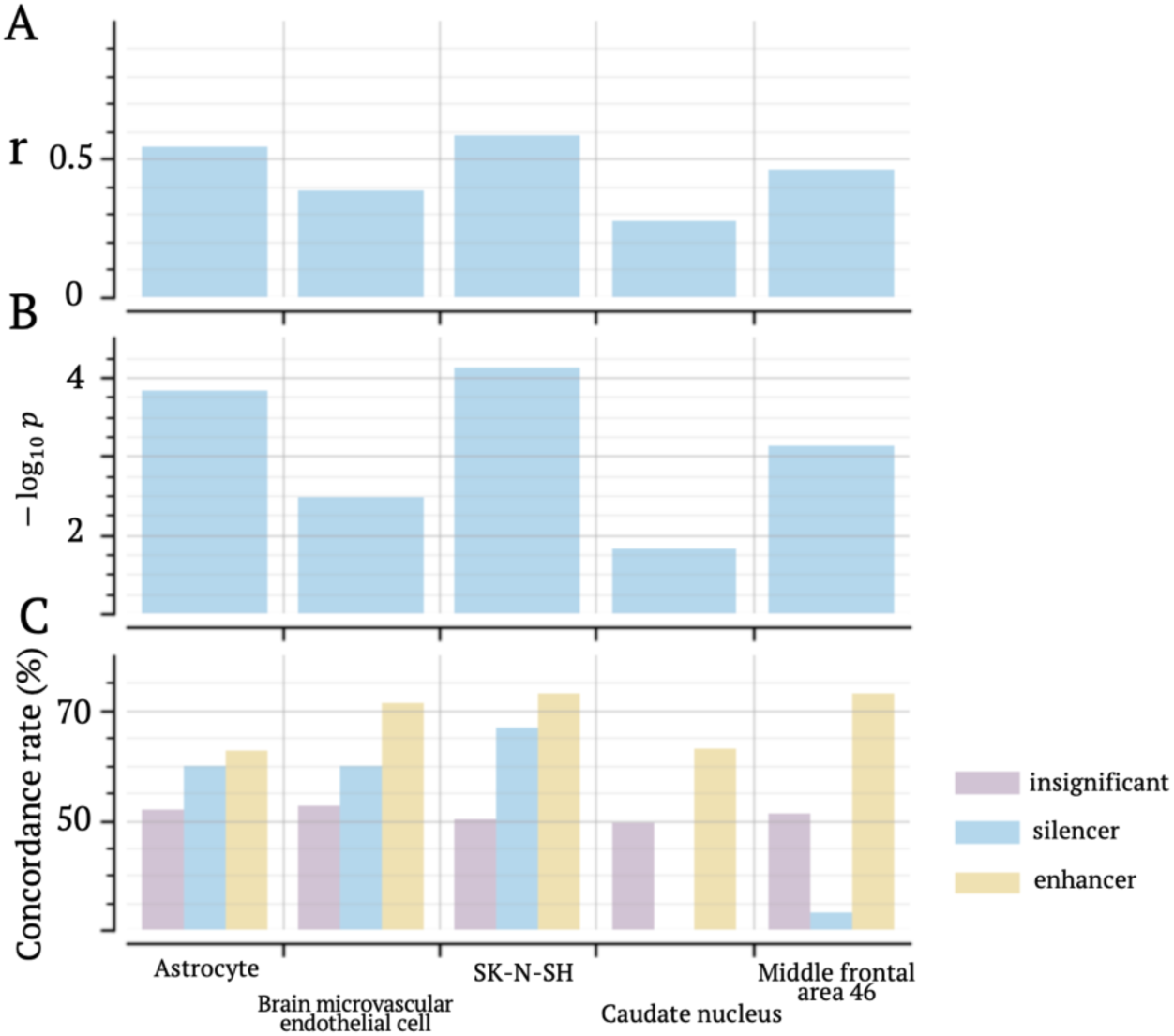
Correlations between Δ*repression* and dementia MPRA scores across CNS biosamples. (A) correlation coefficients between Δ*repression* and dementia MPRA scores. (B) significant p values these ocefficients. (C) concordance rates between Δ*repression* and dementia MPRA scores in three SNP categories: insignificant-Δ*repression* SNPs, significant-Δ*repression* silencer, and enhancer SNPs.

**Figure S22.**
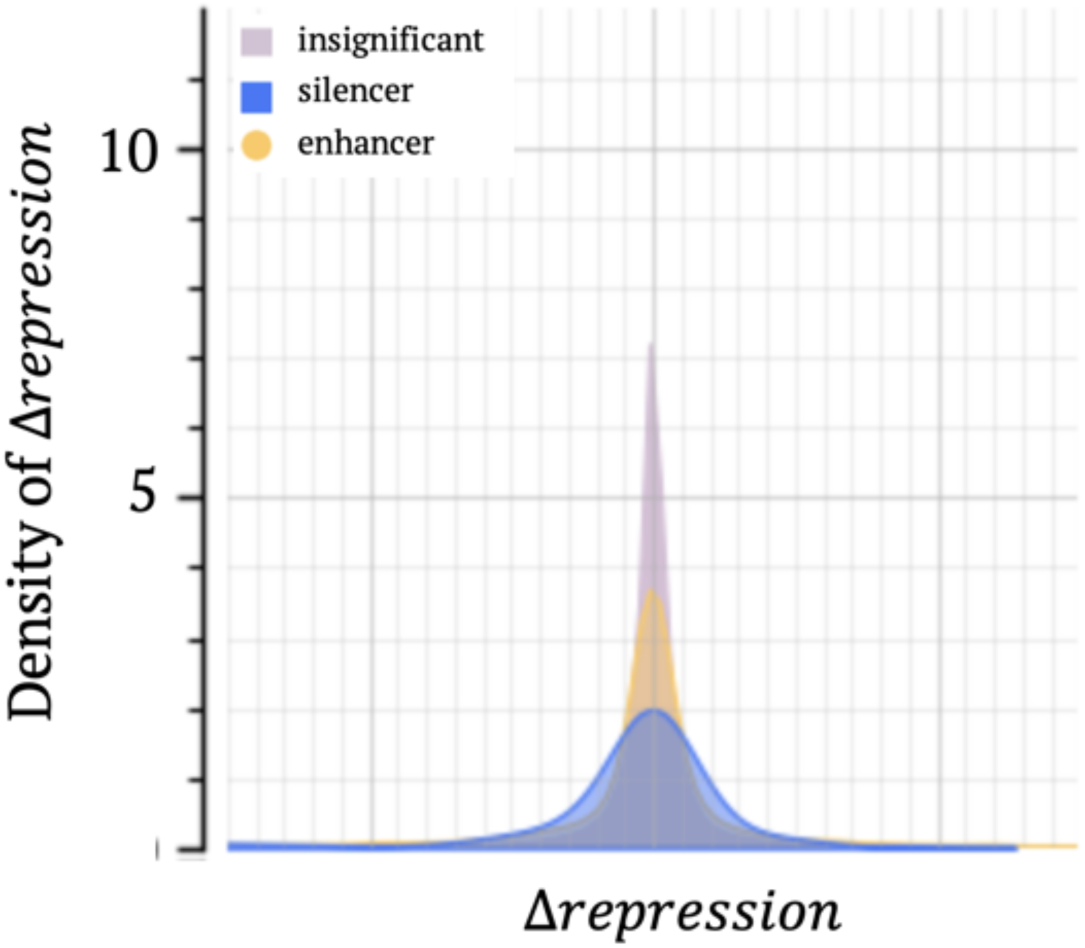
Δ*repression* score distributions for different SNP groups. SNP groups here are those having insignificant mMPRA scores, significant mMPRA scores in candidate silencers, and in candidate enhancers.

**Figure S23.**
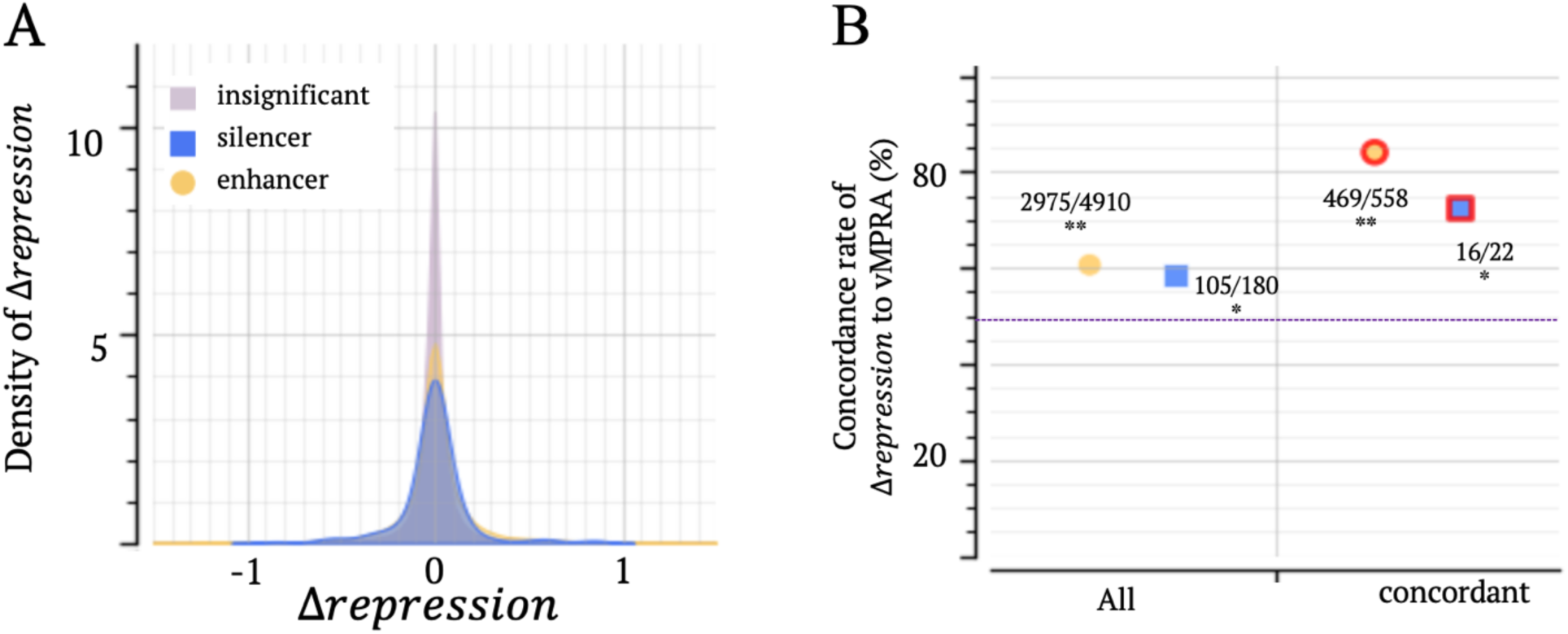
Correlation between Δ*repression* and vMPRA scores. (A) Distribution of Δ*repression* score across SNP groups. SNP groups here are those having insignificant vMPRA scores, significant vMPRA scores in candidate silencers and enhancers. (B) Directional concordance between Δ*repression* and vMPRA scores. “All” represents all significant-vPRA SNPs in candidate silencers or enhancers, while “concordant” represents the SNPs where significant mMPRA and vMPRA scores directionally align. The dash line represents the expectation after randomly shuffling Δ*repression*. ∗∗ *p* < 10^−8^, ∗ *p* < 0.01

**Figure S24.**
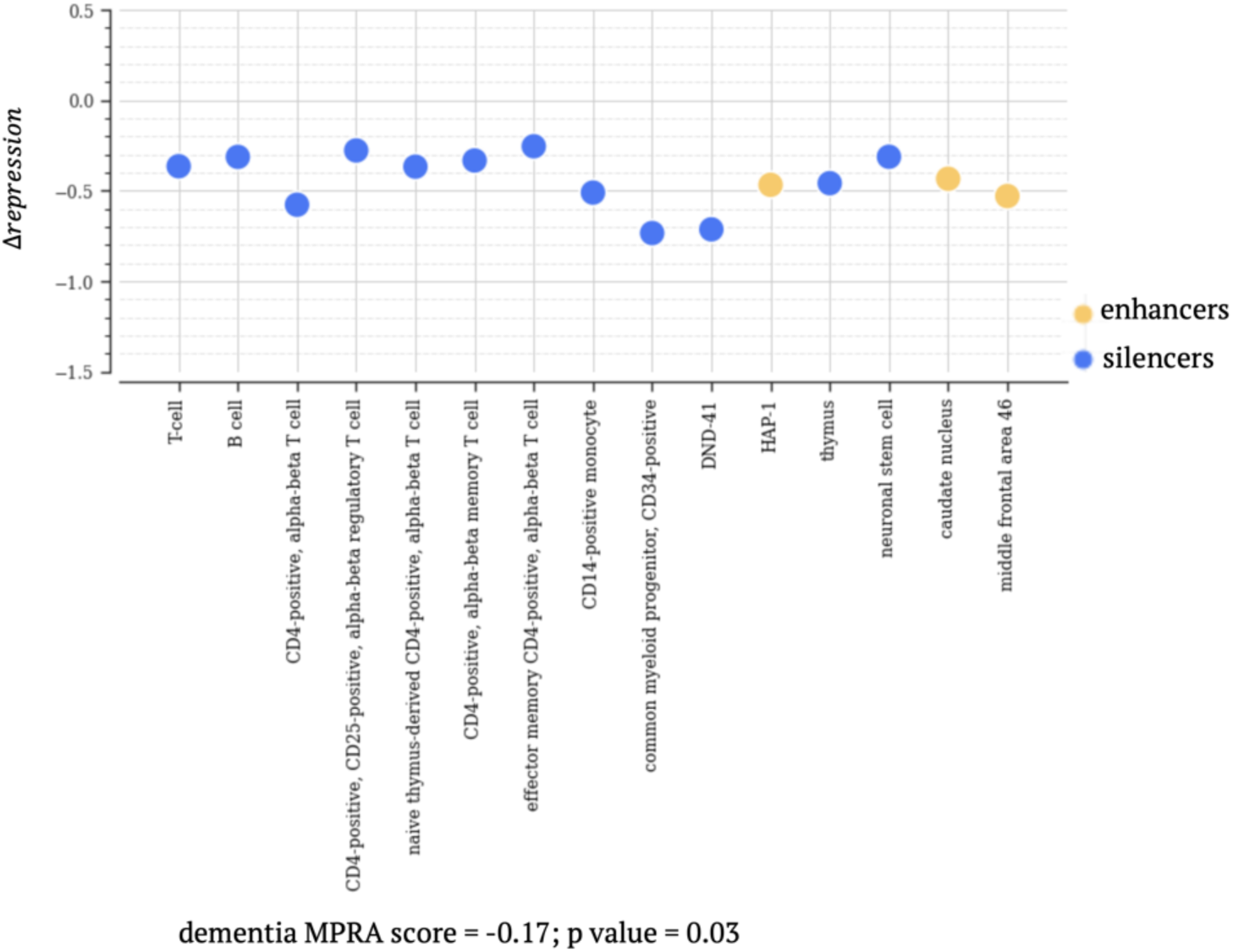
Δ*repression* scores at rs242561. The biosamples where rs242561 is located within a candidate silencer or enhancer are included.

**Table S1**. All files defining training samples for TREDnet models.

**Table S2**. TFBS ChIP-seq files used in this study.

**Table S3**. Enrichment of GWAS SNPs in candidate silencers and enhancers.

**Table S4**. RNA-seq files used in this study.

## Notes

### Competing Interest Statement

The authors have declared no competing interest.

## Reference

1. Abell NS, DeGorter MK, Gloudemans MJ, Greenwald E, Smith KS, He Z, Montgomery SB. 2022. Multiple causal variants underlie genetic associations in humans. Science 375: 1247–1254.

2. Benes FM. 2004. The role of apoptosis in neuronal pathology in schizophrenia and bipolar disorder. Current Opinion in Psychiatry 17.

3. Bento-Pereira C, Dinkova-Kostova AT. 2021. Activation of transcription factor Nrf2 to counteract mitochondrial dysfunction in Parkinson’s disease. Med Res Rev 41: 785–802.

4. Bortolami A, Yu W, Forzisi E, Ercan K, Kadakia R, Murugan M, Fedele D, Estevez I, Boison D, Rasin M-R, Sesti F. 2023. Integrin-KCNB1 potassium channel complexes regulate neocortical neuronal development and are implicated in epilepsy. Cell Death & Differentiation 30: 687–701.

5. Brent JR, Nora B-V, Matthew W, Ecem K, Rosalind R, Gabriele C, Maximilian S, Saba S, Marta C, Josse P et al. 2021. REST Protects Dopaminergic Neurons from Mitochondrial and α-Synuclein Oligomer Pathology in an Alpha Synuclein Overexpressing BAC-Transgenic Mouse Model. The Journal of Neuroscience 41: 3731.

6. Bycroh C, Freeman C, Petkova D, Band G, Elliott LT, Sharp K, Motyer A, Vukcevic D, Delaneau O, O’Connell J et al. 2018. The UK Biobank resource with deep phenotyping and genomic data. Nature 562: 203–209.

7. Casellas A, Mallol C, Salavert A, Jimenez V, Garcia M, Agudo J, Obach M, Haurigot V, Vilà L, Molas M et al. 2015. Insulin-like Growth Factor 2 Overexpression Induces β-Cell Dysfunction and Increases Beta-cell Susceptibility to Damage. J Biol Chem 290: 16772–16785.

8. Claussnitzer M, Cho JH, Collins R, Cox NJ, Dermitzakis ET, Hurles ME, Kathiresan S, Kenny EE, Lindgren CM, MacArthur DG et al. 2020. A brief history of human disease genetics. Nature 577: 179–189.

9. Clihon NE, Hannon E, Harwood JC, Di Florio A, Thomas KL, Holmans PA, Walters JTR, O’Donovan MC, Owen MJ, Pocklington AJ, Hall J. 2019. Dynamic expression of genes associated with schizophrenia and bipolar disorder across development. Translational Psychiatry 9: 74.

10. Clyde D. 2023. Chromatin loops stack up. Nature Reviews Genetics 24: 415–415.

11. Cooper YA, Teyssier N, Dräger NM, Guo Q, Davis JE, Sattler SM, Yang Z, Patel A, Wu S, Kosuri S et al. 2022. Functional regulatory variants implicate distinct transcriptional networks in dementia. Science 377: eabi8654.

12. Della Rosa M, Spivakov M. 2020. Silencers in the spotlight. Nature Genetics 52: 244–245.

13. Doni Jayavelu N, Jajodia A, Mishra A, Hawkins RD. 2020. Candidate silencer elements for the human and mouse genomes. Nature Communications 11: 1061.

14. Du J, Machado-Vieira R, Khairova R. 2011. Synaptic plasticity in the pathophysiology and treatment of bipolar disorder. Curr Top Behav Neurosci 5: 167–185.

15. Dunning AM, Michailidou K, Kuchenbaecker KB, Thompson D, French JD, Beesley J, Healey CS, Kar S, Pooley KA, Lopez-Knowles E et al. 2016. Breast cancer risk variants at 6q25 display different phenotype associations and regulate ESR1, RMND1 and CCDC170. Nat Genet 48: 374–386.

16. Erceg J, Pakozdi T, Marco-Ferreres R, Ghavi-Helm Y, Girardot C, Bracken AP, Furlong EEM. 2017. Dual functionality of cis-regulatory elements as developmental enhancers and Polycomb response elements. Genes & development 31: 590–602.

17. Ernst J, Melnikov A, Zhang X, Wang L, Rogov P, Mikkelsen TS, Kellis M. 2016. Genome-scale high-resolution mapping of activating and repressive nucleotides in regulatory regions. Nature Biotechnology 34: 1180.

18. Farh KK, Marson A, Zhu J, Kleinewieteld M, Housley WJ, Beik S, Shoresh N, Whitton H, Ryan RJ, Shishkin AA et al. 2015. Genetic and epigenetic fine mapping of causal autoimmune disease variants. Nature 518: 337–343.

19. Faurholt-Jepsen M, Frøkjær VG, Nasser A, Jørgensen NR, Kessing LV, Vinberg M. 2021. Associations between the cortisol awakening response and patient-evaluated stress and mood instability in patients with bipolar disorder: an exploratory study. International Journal of Bipolar Disorders 9: 8.

20. Finucane HK, Bulik-Sullivan B, Gusev A, Trynka G, Reshef Y, Loh P-R, Anwla V, Xu H, Zang C, Farh K et al. 2015. Partitioning heritability by functional annotation using genome-wide association summary statistics. Nature Genetics 47: 1228–1235.

21. Fulco CP, Nasser J, Jones TR, Munson G, Bergman DT, Subramanian V, Grossman SR, Anyoha R, Doughty BR, Patwardhan TA et al. 2019. Activity-by-contact model of enhancer-promoter regulation from thousands of CRISPR perturbations. Nat Genet 51: 1664–1669.

22. Grant CE, Bailey TL, Noble WS. 2011. FIMO: scanning for occurrences of a given motif. Bioinformatics 27: 1017–1018.

23. Grass JA, Boyer ME, Pal S, Wu J, Weiss MJ, Bresnick EH. 2003. GATA-1-dependent transcriptional repression of GATA-2 via disruption of positive autoregulation and domain-wide chromatin remodeling. Proceedings of the National Academy of Sciences 100: 8811–8816.

24. Hansen TJ, Hodges E. 2022. ATAC-STARR-seq reveals transcription factor-bound activators and silencers across the chromatin accessible human genome. Genome Research doi:10.1101/gr.276766.122.

25. Huang D, Ovcharenko I. 2022. Enhancer–silencer transitions in the human genome. Genome Research 32: 437–448.

26. Huang D, Petrykowska HM, Miller BF, Elnitski L, Ovcharenko I. 2019. Identification of human silencers by correlating cross-tissue epigenetic profiles and gene expression. Genome Research.

27. Hudaiberdiev S, Taylor DL, Song W, Narisu N, Bhuiyan RM, Taylor HJ, Tang X, Yan T, Swih AJ, Bonnycastle LL et al. 2023. Modeling islet enhancers using deep learning identifies candidate causal variants at loci associated with T2D and glycemic traits. Proc Natl Acad Sci U S A 120: e2206612120.

28. Hussain S, Sadouni N, van Essen D, Dao LTM, Ferré Q, Charbonnier G, Torres M, Gallardo F, Lecellier C-H, Sexton T et al. 2023. Short tandem repeats are important contributors to silencer elements in T cells. Nucleic Acids Research 51: 4845–4866.

29. Jahangir M, Zhou JS, Lang B, Wang XP. 2021. GABAergic System Dysfunction and Challenges in Schizophrenia Research. Front Cell Dev Biol 9: 663854.

30. Kemp DE, Gao K, Chan PK, Ganocy SJ, Findling RL, Calabrese JR. 2010. Medical comorbidity in bipolar disorder: relationship between illnesses of the endocrine/metabolic system and treatment outcome. Bipolar Disord 12: 404–413.

31. Konrad EDH, Nardini N, Caliebe A, Nagel I, Young D, Horvath G, Santoro SL, Shuss C, Ziegler A, Bonneau D et al. 2019. CTCF variants in 39individuals with a variable neurodevelopmental disorder broaden the mutational andclinical spectrum. Genetics in Medicine 21: 2723–2733.

32. Landrum MJ, Chitipiralla S, Brown GR, Chen C, Gu B, Hart J, Hoffman D, Jang W, Kaur K, Liu C, et al. 2020. ClinVar: improvements to accessing data. Nucleic Acids Research 48: D835–D844.

33. Lee SH, Ripke S, Neale BM, Faraone SV, Purcell SM, Perlis RH, Mowry BJ, Thapar A, Goddard ME, Witte JS et al. 2013. Genetic relationship between five psychiatric disorders estimated from genome-wide SNPs. Nat Genet 45: 984–994.

34. Li P, Spolski R, Liao W, Wang L, Murphy TL, Murphy KM, Leonard WJ. 2012. BATF-JUN is critical for IRF4-mediated transcription in T cells. Nature 490: 543–546.

35. Li Q, Verma IM. 2002. NF-κB regulation in the immune system. Nature Reviews Immunology 2: 725–734.

36. Li W, Fu X, Liu R, Wu C, Bai J, Xu Y, Zhao Y, Xu Y. 2013. FOXC2 often overexpressed in glioblastoma enhances proliferation and invasion in glioblastoma cells. Oncol Res 21: 111–120.

37. Li Y, Ma C, Li S, Wang J, Li W, Yang Y, Li X, Liu J, Yang J, Liu Y et al. 2022. Regulatory Variant rs2535629 in ITIH3 Intron Confers Schizophrenia Risk By Regulating CTCF Binding and SFMBT1 Expression. Adv Sci (Weinh) 9: e2104786.

38. Li Z, Gao E, Zhou J, Han W, Xu X, Gao X. 2023. Applications of deep learning in understanding gene regulation. Cell Reports Methods 3: 100384.

39. Maatouk L, Compagnion AC, Sauvage MC, Bemelmans AP, Leclere-Turbant S, Cirotteau V, Tohme M, Beke A, Trichet M, Bazin V et al. 2018. TLR9 activation via microglial glucocorticoid receptors contributes to degeneration of midbrain dopamine neurons. Nat Commun 9: 2450.

40. Malone J, Holloway E, Adamusiak T, Kapushesky M, Zheng J, Kolesnikov N, Zhukova A, Brazma A, Parkinson H. 2010. Modeling sample variables with an Experimental Factor Ontology. Bioinformatics 26: 1112–1118.

41. Marxreiter F, Regensburger M, Winkler J. 2013. Adult neurogenesis in Parkinson’s disease. Cell Mol Life Sci 70: 459–473.

42. Maurano MT, Humbert R, Rynes E, Thurman RE, Haugen E, Wang H, Reynolds AP, Sandstrom R, Qu H, Brody J et al. 2012. Systematic localization of common disease-associated variation in regulatory DNA. Science 337: 1190–1195.

43. McLean CY, Bristor D, Hiller M, Clarke SL, Schaar BT, Lowe CB, Wenger AM, Bejerano G. 2010. GREAT improves functional interpretation of cis-regulatory regions. Nature Biotechnology 28: 495–501.

44. McMahon A, Malangone C, Suveges D, Sollis E, Cunningham F, Riat HS, MacArthur JA L, Hayhurst J, Morales J, Guillen JA et al. 2018. The NHGRI-EBI GWAS Catalog of published genome-wide association studies, targeted arrays and summary statistics 2019. Nucleic Acids Research 47: D1005–D1012.

45. Moons R, Konijnenberg A, Mensch C, Van Elzen R, Johannessen C, Maudsley S, Lambeir A-M, Sobott F. 2020. Metal ions shape α-synuclein. Scientific Reports 10: 16293.

46. Mouri K, Dewey HB, Castro R, Berenzy D, Kales S, Tewhey R. 2023. Whole-genome functional characterization of RE1 silencers using a modified massively parallel reporter assay. Cell Genomics 3: 100234.

47. Nakazawa K, Zsiros V, Jiang Z, Nakao K, Kolata S, Zhang S, Belforte JE. 2012. GABAergic interneuron origin of schizophrenia pathophysiology. Neuropharmacology 62: 1574–1583.

48. Ngan CY, Wong CH, Tjong H, Wang W, Goldfeder RL, Choi C, He H, Gong L, Lin J, Urban B et al. 2020. Chromatin interaction analyses elucidate the roles of PRC2-bound silencers in mouse development. Nature Genetics 52: 264–272.

49. Pang B, Snyder MP. 2020. Systematic identification of silencers in human cells. Nature Genetics 52: 254–263.

50. Quinonez SC, Innis JW. 2014. Human HOX gene disorders. Molecular Genetics and Metabolism 111: 4–15.

51. Ruderfer DM, Ripke S, McQuillin A, Boocock J, Stahl EA, Pavlides JMW, Mullins N, Charney AW, Ori APS, Loohuis LMO et al. 2018. Genomic Dissection of Bipolar Disorder and Schizophrenia, Including 28 Subphenotypes. Cell 173: 1705–1715.e1716.

52. Salameh TJ, Wang X, Song F, Zhang B, Wright SM, Khunsriraksakul C, Ruan Y, Yue F. 2020. A supervised learning framework for chromatin loop detection in genome-wide contact maps. Nature Communications 11: 3428.

53. Santarius T, Shipley J, Brewer D, Stratton MR, Cooper CS. 2010. A census of amplified and overexpressed human cancer genes. Nature Reviews Cancer 10: 59–64.

54. Schaniel C, Ang YS, Ratnakumar K, Cormier C, James T, Bernstein E, Lemischka IR, Paddison PJ. 2009. Smarcc1/Baf155 couples self-renewal gene repression with changes in chromatin structure in mouse embryonic stem cells. Stem Cells 27: 2979–2991.

55. Severance EG, Dickerson F, Yolken RH. 2020. Complex Gastrointestinal and Endocrine Sources of Inflammation in Schizophrenia. Front Psychiatry 11: 549.

56. Sherman BT, Hao M, Qiu J, Jiao X, Baseler MW, Lane HC, Imamichi T, Chang W. 2022. DAVID: a web server for functional enrichment analysis and functional annotation of gene lists (2021 update). Nucleic Acids Res 50: W216–221.

57. Sherry ST, Ward MH, Kholodov M, Baker J, Phan L, Smigielski EM, Sirotkin K. 2001. dbSNP: the NCBI database of genetic variation. Nucleic Acids Res 29: 308–311.

58. Steck AK, Rewers MJ. 2011. Genetics of type 1 diabetes. Clin Chem 57: 176–185.

59. Tenney AP, Di Gioia SA, Webb BD, Chan W-M, de Boer E, Garnai SJ, Barry BJ, Ray T, Kosicki M, Robson CD et al. 2023. Noncoding variants alter GATA2 expression in rhombomere 4 motor neurons and cause dominant hereditary congenital facial paresis. Nature Genetics 55: 1149–1163.

60. Tewhey R, Kotliar D, Park DS, Liu B, Winnicki S, Reilly SK, Andersen KG, Mikkelsen TS, Lander ES, Schaffner SF, Sabeti PC. 2016. Direct Identification of Hundreds of Expression-Modulating Variants using a Multiplexed Reporter Assay. Cell 165: 1519–1529.

61. The Alliance of Genome Resources C. 2020. Alliance of Genome Resources Portal: unified model organism research platorm. Nucleic Acids Research 48: D650–D658.

62. The GTEx Consortium. 2015. The Genotype-Tissue Expression (GTEx) pilot analysis: Multitissue gene regulation in humans. Science 348: 648–660.

63. van Arensbergen J, Pagie L, FitzPatrick VD, de Haas M, Baltissen MP, Comoglio F, van der Weide RH, Teunissen H, Võsa U, Franke L et al. 2019. High-throughput identification of human SNPs affecting regulatory element activity. Nature Genetics 51: 1160–1169.

64. Villar D, Berthelot C, Aldridge S, Rayner Tim F, Lukk M, Pignatelli M, Park Thomas J, Deaville R, Erichsen Jonathan T, Jasinska Anna J et al. 2015. Enhancer Evolution across 20 Mammalian Species. Cell 160: 554–566.

65. Watanabe K, Stringer S, Frei O, Umićević Mirkov M, de Leeuw C, Polderman TJC, van der Sluis S, Andreassen OA, Neale BM, Posthuma D. 2019. A global overview of pleiotropy and genetic architecture in complex traits. Nature Genetics 51: 1339–1348.

66. Xiusheng Z, Chao W, Dashuai K, Jing L, Biao D, Yi G, Siyuan K, Lei H, Yuwen L, Yubo Z. 2023. Genome-wide identification of silencers in human cells. bioRxiv doi:10.1101/2023.06.20.545673: 2023.2006.2020.545673.

67. Yan J, Qiu Y, Ribeiro Dos Santos AM, Yin Y, Li YE, Vinckier N, Nariai N, Benaglio P, Raman A, Li X et al. 2021. Systematic analysis of binding of transcription factors to noncoding variants. Nature 591: 147–151.

68. Yoshifuji H, Terao C. 2020. Roles of cytotoxic lymphocytes and MIC/LILR families in pathophysiology of Takayasu arteritis. Inflammation and Regeneration 40: 9.

69. Young FI, Keruzore M, Nan X, Gennet N, Bellefroid EJ, Li M. 2017. The doublesex-related Dmrta2 safeguards neural progenitor maintenance involving transcriptional regulation of Hes1. Proc Natl Acad Sci U S A 114: E5599–e5607.

70. Yu WP, Yew K, Rajasegaran V, Venkatesh B. 2007. Sequencing and comparative analysis of fugu protocadherin clusters reveal diversity of protocadherin genes among teleosts. BMC Evol Biol 7: 49.

71. Zhang F, Lupski JR. 2015. Non-coding genetic variants in human disease. Hum Mol Genet 24: R102–110.

72. Zhang J, Baran J, Cros A, Guberman JM, Haider S, Hsu J, Liang Y, Rivkin E, Wang J, Whitty B et al. 2011. International Cancer Genome Consortium Data Portal--a one-stop shop for cancer genomics data. Database: the journal of biological databases and curation 2011: bar026–bar026.

73. Zhou J, Theesfeld CL, Yao K, Chen KM, Wong AK, Troyanskaya OG. 2018. Deep learning sequence-based ab initio prediction of variant effects on expression and disease risk. Nature Genetics 50: 1171–1179.

74. Zhu X, Wang C, Kong D, Luo J, Deng B, Gu Y, Kong S, Huang L, Liu Y, Zhang Y. 2023. Genome-wide identification of silencers in human cells. bioRxiv doi:10.1101/2023.06.20.545673: 2023.2006.2020.545673.

## References

1 Luo, Y., et al. New developments on the Encyclopedia of DNA Elements (ENCODE) data portal. Nucleic Acids Res 48, D882–d889 (2020). 10.1093/nar/gkz1062

2 Frankish, A. et al. GENCODE 2021. Nucleic Acids Research 49, D916–D923 (2021). 10.1093/nar/gkaa1087

3 The GTEx Consortium. The Genotype-Tissue Expression (GTEx) pilot analysis: Multitissue gene regulation in humans. Science 348, 648–660 (2015). 10.1126/science.1262110

